# Estimating probabilities of malaria importation in southern Mozambique through *P. falciparum* genomics and mobility patterns

**DOI:** 10.1101/2025.05.01.25326793

**Authors:** Arnau Pujol, Arlindo Chidimatembue, Clemente da Silva, Simone Boene, Henriques Mbebe, Pau Cisteró, Carla García-Fernández, Arnau Vañó-Boira, Dário Tembisse, José Inácio, Glória Matambisso, Fabião Luis, Nelo Ndimande, Humberto Munguambe, Lidia Nhamussua, Wilson Simone, Andrés Aranda-Díaz, Manuel García-Ulloa, Neide Canana, Maria Tusell, Júlia Montaña, Laura Fuente-Soro, Khalid Ussene Bapu, Maxwell Murphy, Bernardete Rafael, Eduard Rovira-Vallbona, Caterina Guinovart, Bryan Greenhouse, Sonia Maria Enosse, Francisco Saúte, Pedro Aide, Baltazar Candrinho, Alfredo Mayor

## Abstract

**Background:** Imported malaria is a critical obstacle to achieving elimination in low transmission settings. Characterising malaria importation and transmission sources using human mobility and parasite genomics has the potential to inform elimination strategies, but tools combining both types of data are lacking.

**Methods:** We developed a novel Bayesian approach that provides individual importation probabilities and geographic origin of *P. falciparum* cases by combining epidemiological, human mobility and parasite genetic data. Spatial genetic structure and connectivity were assessed using microhaplotype-based genetic relatedness (identity-by-descent) from 1467 *P. falciparum* samples collected from 9 provinces in Mozambique during 2022, including 200 samples from two very-low transmission elimination-targeted districts (Magude and Matutuine) in the south. Travel reports were combined with genetic relatedness metrics to classify clinical cases as local or imported.

**Results:** Genetic relatedness between parasites from southern and northern/central Mozambique was lower (0.017) than average (0.024, p<0.001). 43.5% (87/200) of infections in elimination-targeted districts were classified as imported, had a higher genetic complexity (OR=1.4, CI=[1.0, 1.9], p=0.038) compared to local cases and originated mainly from Inhambane (62% [54/87]). Significant differences in the odds of a case being imported were found between the two study districts (OR=6.6, CI=[2.3,25.4], p<0.001), with Magude district (11.1%, 3/27) showing lower importation rates than Matutuine (48.6%, 84/173) district.

**Conclusions:** Differences in importation rates observed between both elimination districts suggest the need for fine-scale analysis to tailor cost-effective elimination strategies. Importation is strongly impacting malaria incidence in Matutuine district, and increasing efforts to reduce malaria burden in their sources of transmission (especially in Inhambane province), as well as targeting travelers to central and northern Mozambique, could significantly contribute to malaria elimination in the south.

**Funding:** Bill and Melinda Gates Foundation (INV-019032, INV-067310), the European Union’s Horizon 2020 research and innovation programme (Marie Skłodowska-Curie grant 890477), “laCaixa” Foundation (ID 100010434, fellowship code LCF/BQ/PR24/12050009).

## Introduction

Despite the efforts during the last decades, malaria elimination remains challenging, especially in Sub-Saharan Africa^1–6^. Detection, characterisation and monitoring of malaria infections are key to control and elimination^7^. However, decisions become more complex in low transmission areas given the heterogeneity in transmission patterns, requiring better tools to identify the key sources of transmission^8^.

One of the key challenges for malaria elimination in very-low transmission settings is parasite importation, which can sustain transmission during “last mile” efforts^9–14^. Identifying imported cases and their transmission sources, together with associated risk factors, is key to improving targeted efforts for malaria elimination in very-low transmission areas. Previous studies that aimed to classify reported cases as local or imported used epidemiological data based on travel reports (assuming or modelling that the infections occurred during the trip)^15–18^, or mobile phone data and geospatial modelling to characterise population mobility and infer the potential impact of importation^19–22^. Other studies used parasite genomics to assess the spatial connectivity of genetic populations to infer transmission flow, migration patterns or infection origin^23–37^. However, a study comparing mobility, phone and parasite genetic data brought distinct conclusions on the spatial connectivities due to intrinsic biases of the different data sources^30^. Two studies which combined mobility (travel reports and mobile phone data) with genetic data found a positive association between mobility and parasite genetic relatedness, providing evidence of importation^21,30^. However, none of these studies combined mobility and genetic data simultaneously to provide importation rates in the populations or for individual case classification.

We present a new method to provide case classification as individual importation probabilities by combining travel, epidemiological, and *P. falciparum* genetic data. We apply this method in the context of Mozambique, a country with medium-high malaria transmission in central and northern Mozambique and low transmission levels in the south. In particular, we study the role of importation in two southern districts from Maputo province, Magude and Matutuine. Magude is a mainly rural interior district, bordering the National Kruger Park (South Africa), while Matutuine is a district close to Maputo city, with better connection and communication infrastructures and bordering South Africa from the south. The study uses data from children (a common sentinel group for symptomatic cases and surveillance) from 9 (out of 11) provinces of Mozambique to assess the spatial structure (dependence on pairwise geographical distance) and differentiation (across areas) of *P. falciparum* genetic populations in the country, quantify the levels of importation in the very low transmission districts of Magude and Matutuine in southern Mozambique, and identify sources of transmission and risk factors associated with human mobility and malaria importation.

## Results

### Participant recruitment, sample and data collection

The study was conducted during 2022 in 7 health facilities (HF) of Magude and 13 HF of Matutuine districts, both in Maputo province (**Figure 1**). These are very-low transmission areas (with less than 52 yearly cases per 1,000 people in all HFs) with the highest rainfall between January and May. A total of 809 *P. falciparum* positive clinical cases were reported, from which 609 (75.3%, 609/809) rapid diagnostic tests (RDTs) were available for parasite sequencing and 540 (66.7%, 540/809) were sequenced successfully (with allele calls passing both negative controls and allele frequency filters, see Methods). Demographic data and travel reports were available for 232 (28.7%, 232/809) of the samples, and 200 (24.7%, 200/809) resided in the area and passed sequencing coverage and depth requirements (at least 123 loci with >100 reads, see Methods section for more details)^38,39^ (**Table 1**, **Figure 2**). From these 200, 52.5% (105/200) reported a trip during the last 28 days, with 54.3% (57/105) of those having travelled to Inhambane province, the main travel destination (**Figure 3**, **Table 2**). Travels were represented geographically in **Figure 3B**. The other principal destinations were Zambézia (14.3%, 15/105), Gaza (12.4%, 13/105) and Maputo provinces (9.5%, 10/105). Significant differences were found between clinical cases from Magude and Matutuine districts with reported travel, with those from Matutuine showing higher travel rates (11.1% [3/27] in Magude versus 59.0% [102/173] in Matutuine, p<0.001), and occupation (p=0.021), but not on season (p=0.066), age (p=0.473), sex (p=0.194) or travel destination (p=0.374) (**Table 1**).

**Figure. 1:**
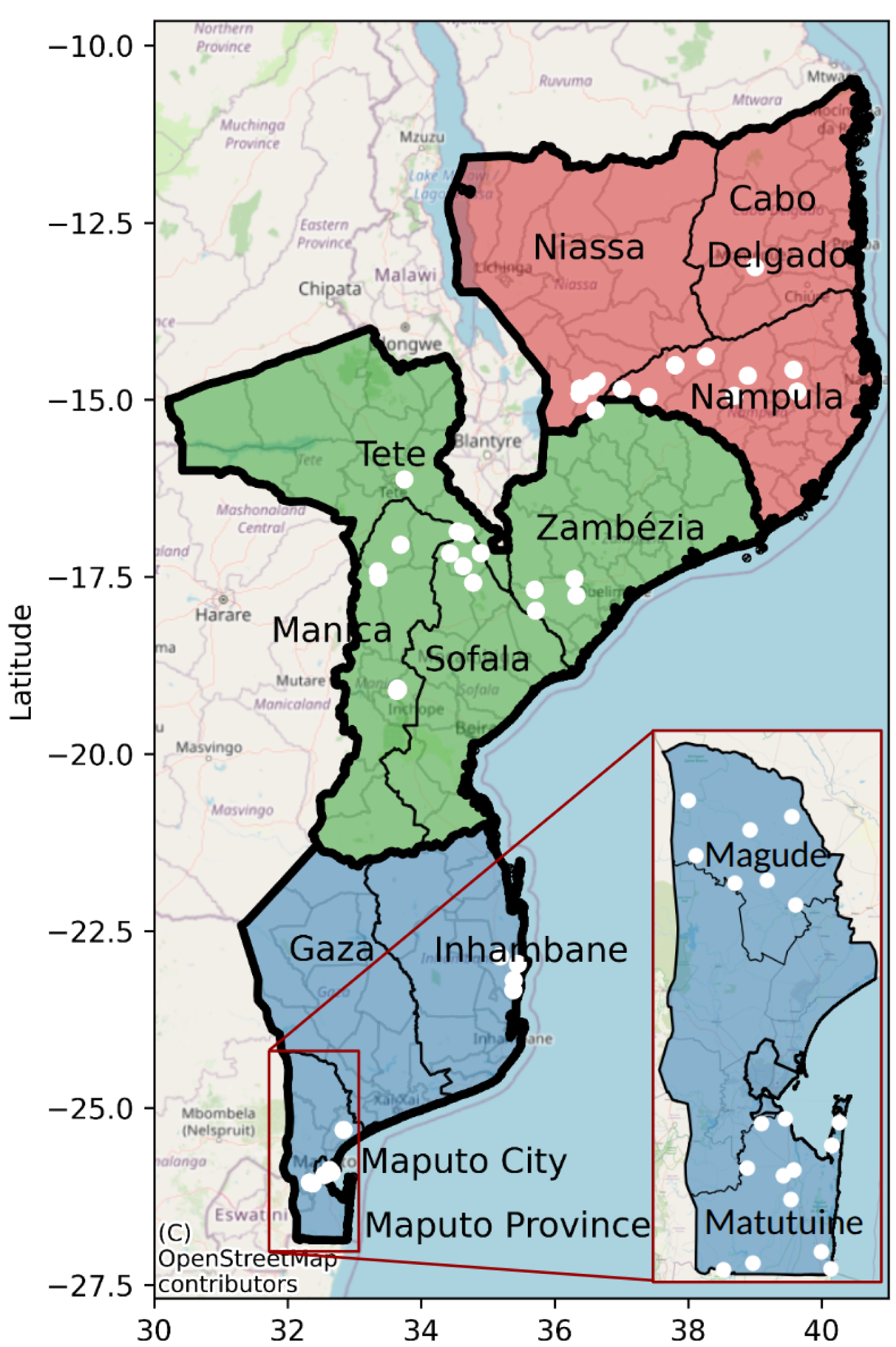
Map of the study health facilities. Provinces were colours according to their region: north (red), centre (green) or south (blue). White dots show the locations of the health facilities included in the study, with a zoom in to show the ones included in Magude and Matutuine districts. Maps used OpenStreetMap data, available under the Open Database License.

**Figure 2:**
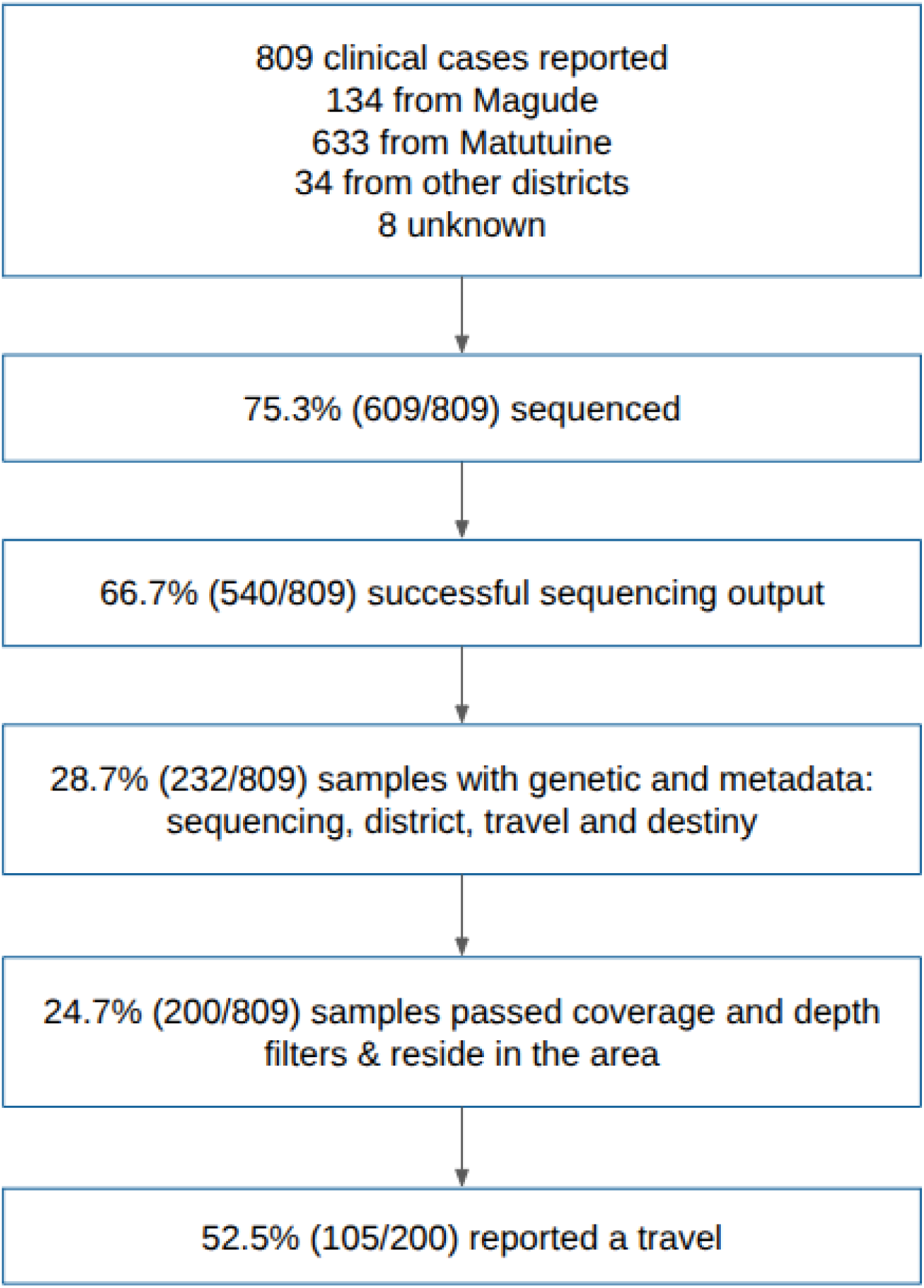
Flowchart of the P. falciparum samples and data from Magude and Matutuine districts collected in 2022.

**Figure 3:**
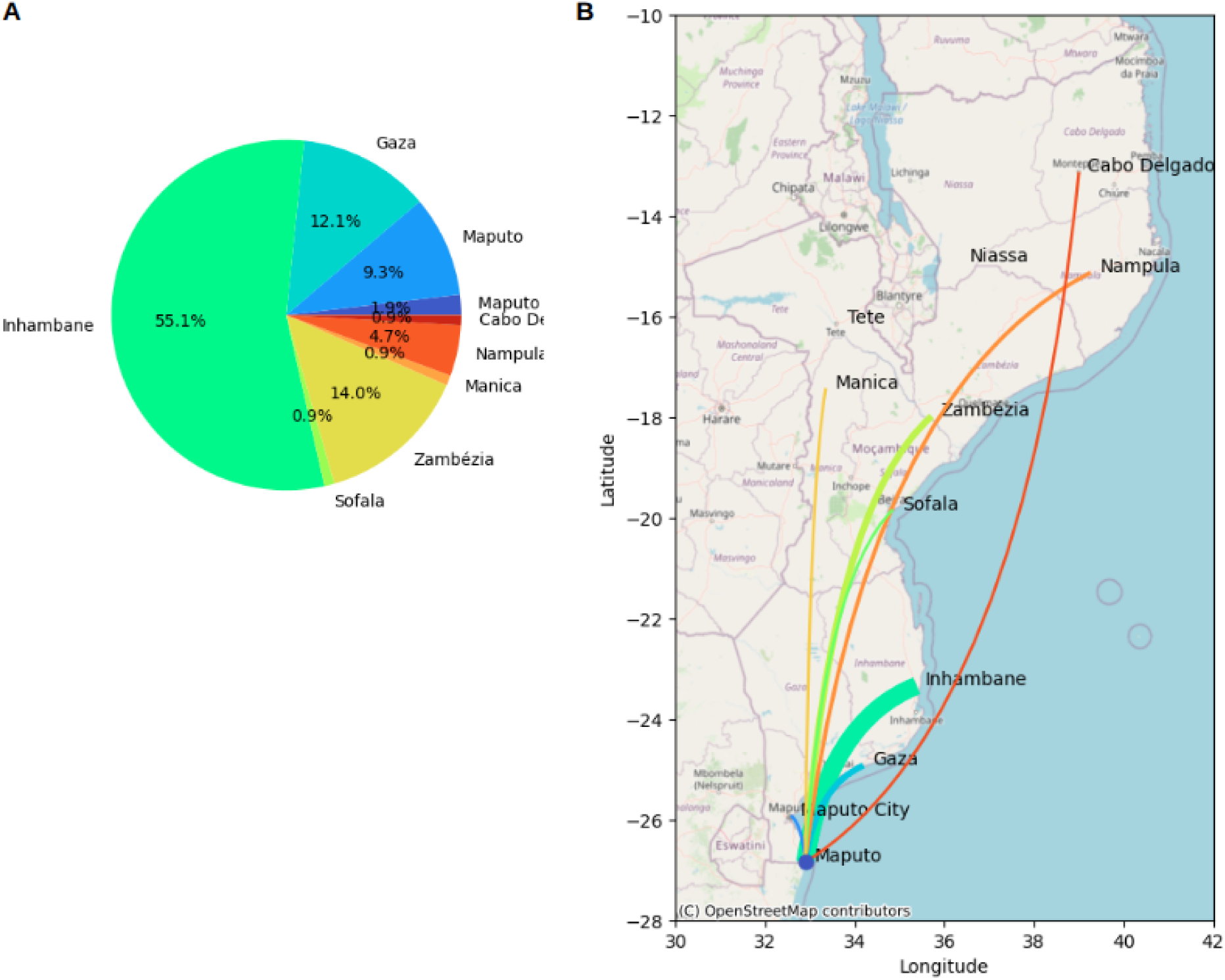
Statistics of travel reports. **A)** Pie chart showing the distributions of the travel destination provinces in sampled individuals from Maputo province (Magude and Matutuine). Colours show the provinces from blue (south) to red (north). **B)** Spatial connectivity based on travel history. Line widths are proportional to the number of travels reported from Maputo province to their destination province, with the same colors as in A. Maps used OpenStreetMap data, available under the Open Database License.

**Table 1:** Sample size and characteristics of the P*. falciparum* clinical cases in Magude and Matutuine with travel history data.

| Category |  | Magude | Matutuine | p-value (Chi2) | p-value (T-test) |
| --- | --- | --- | --- | --- | --- |
| **N** |  | 27 | 173 | NA | NA |
| **Dry season (%)** |  | 4 (14.8) | 60 (34.7) | 0.066 |  |
| **Mean age (SD)** |  | 21.56 (14.1) | 24.12 (17.6) |  | 0.473 |
| **Female (%)** |  | 10 (37.0) | 91 (52.6) | 0.194 |  |
| **Occupation, n (%)** | Domestic | 3 (11.1) | 41 (23.7) | 0.021 |  |
|  | Student | 9 (33.3) | 16 (9.2) |  |  |
|  | Agriculture | 3 (11.1) | 19 (11.0) |  |  |
|  | Other | 11 (40.7) | 47 (27.2) |  |  |
|  | Unemployed | 1 (3.7) | 50 (28.9) |  |  |
| **Reported travel within previous 28 days (%)** |  | 3 (11.1) | 102 (59.0) | <0.001 |  |
| **Province travel destination** | Gaza | 2 | 11 | 0.374 |  |
|  | Inhambane | 1 | 56 |  |  |
|  | Maputo | 0 | 10 |  |  |
|  | Zambézia | 0 | 15 |  |  |
|  | Manica | 0 | 1 |  |  |
|  | Maputo City | 0 | 2 |  |  |
|  | Sofala | 0 | 1 |  |  |
|  | Nampula | 0 | 5 |  |  |
|  | Cabo Delgado | 0 | 1 |  |  |

**Table 2:** Travel destinations among the 105 *P. falciparum* clinical cases in Magude and Matutuine that reported having travelled during the previous 28 days. Provinces of the travel destinations reported (left column) and the total number of travels (and percentage) reported to each province (right column).

| Travel destination province | Number of travels n (%) |
| --- | --- |
| Maputo City | 2 (1.9%) |
| Maputo Province | 10 (9.5%) |
| Gaza | 13 (12.4%) |
| Inhambane | 57 (54.3%) |
| Sofala | 1 (1.0%) |
| Manica | 1 (1.0%) |
| Zambézia | 15 (14.3%) |
| Nampula | 5 (4.8%) |
| Cabo Delgado | 1 (1.0%) |
| Total | 105 |

Additionally, 949 dried blood spot samples from *P. falciparum* uncomplicated clinical malaria cases were collected in 9 provinces during the 2022 rainy season, in the context of annual health facility surveys (HFS) or clinical trials^40^ (**Figure 1**, **Table 3**). The number of samples sequenced per province ranged between 44 and 95, with the exception of Inhambane province which included 345 samples for a deeper spatial analysis in this province (**Table 3**). Significant differences between provinces were found in terms of cases’ age (p<0.001), but not in gender (p=0.442). Analysis were conducted both at province and regional level, where provinces were classified as follows: south (Maputo, Inhambane), centre (Sofala, Manica, Tete, Zambézia) and north (Nampula, Niassa, Cabo Delgado).

**Table 3:** Convenience sampling from selected health facilities in 9 provinces: number of samples and patient characteristics. Province: province of the health facilities from where the samples were collected. N: number of samples collected in each province. Mean age (SD): mean and standard deviation of the participant ages from each province. Female fraction (%): fraction of females of the participants from each province.

| Province | N | Mean age (SD) | Female fraction (%) |
| --- | --- | --- | --- |
| Maputo | 95 | 20.55 (18.78) | 47.87 |
| Inhambane | 345 | 6.17 (2.67) | 54.44 |
| Sofala | 80 | 5.5 (2.4) | 50 |
| Manica | 65 | 5.55 (2.34) | 47.69 |
| Tete | 83 | 2.3 (1.49) | 43.9 |
| Zambézia | 72 | 4.55 (2.17) | 56.94 |
| Nampula | 44 | 1.45 (0.91) | 43.18 |
| Niassa | 79 | 5.68 (2.68) | 43.04 |
| Cabo Delgado | 86 | 2.22 (1.28) | 51.16 |
| p-value (*=T-test, **=Chi-square test) |  | <0.001* | 0.442** |

### Country-wide spatial trends of *P. falciparum* genetic relatedness

DNA extracted from DBS or RDTs was sequenced using the MAD^4^HatTeR targeted amplicon sequencing panel and 165 microhaplotype loci were used to calculate diversity metrics^41^. A significant spatial pattern of genetic relatedness (*R*, defined as the fraction of related (identity-by-descent, IBD) pairs (IBD>0.1 and p<0.05), see Methods) in *P. falciparum* populations within and across provinces was found (**Figure 4A**, with a geographical representation of the results in **Figure 4B**) with a strong south-centre/north differentiation (**Figure 4C,D**). Sample pairs within centre/north Mozambique presented a higher *R* (0.028, `7276`/`258572` pairs) than the average across regions (0.0238, p<0.001). However, *R* between south and centre/north sample pairs was lower than average (0.0174, 1950/111980 pairs, p<0.001) (**Figures 4,5**). The highest *R* from Maputo province was with Nampula (*R*=0.029 [126/4400 pairs]). When stratifying Maputo and Inhambane provinces by district (samples from these provinces were collected in two districts), *R* was not significantly different within Maputo districts than across districts from different provinces (**Figures 4E,5D**).

**Figure 4:**
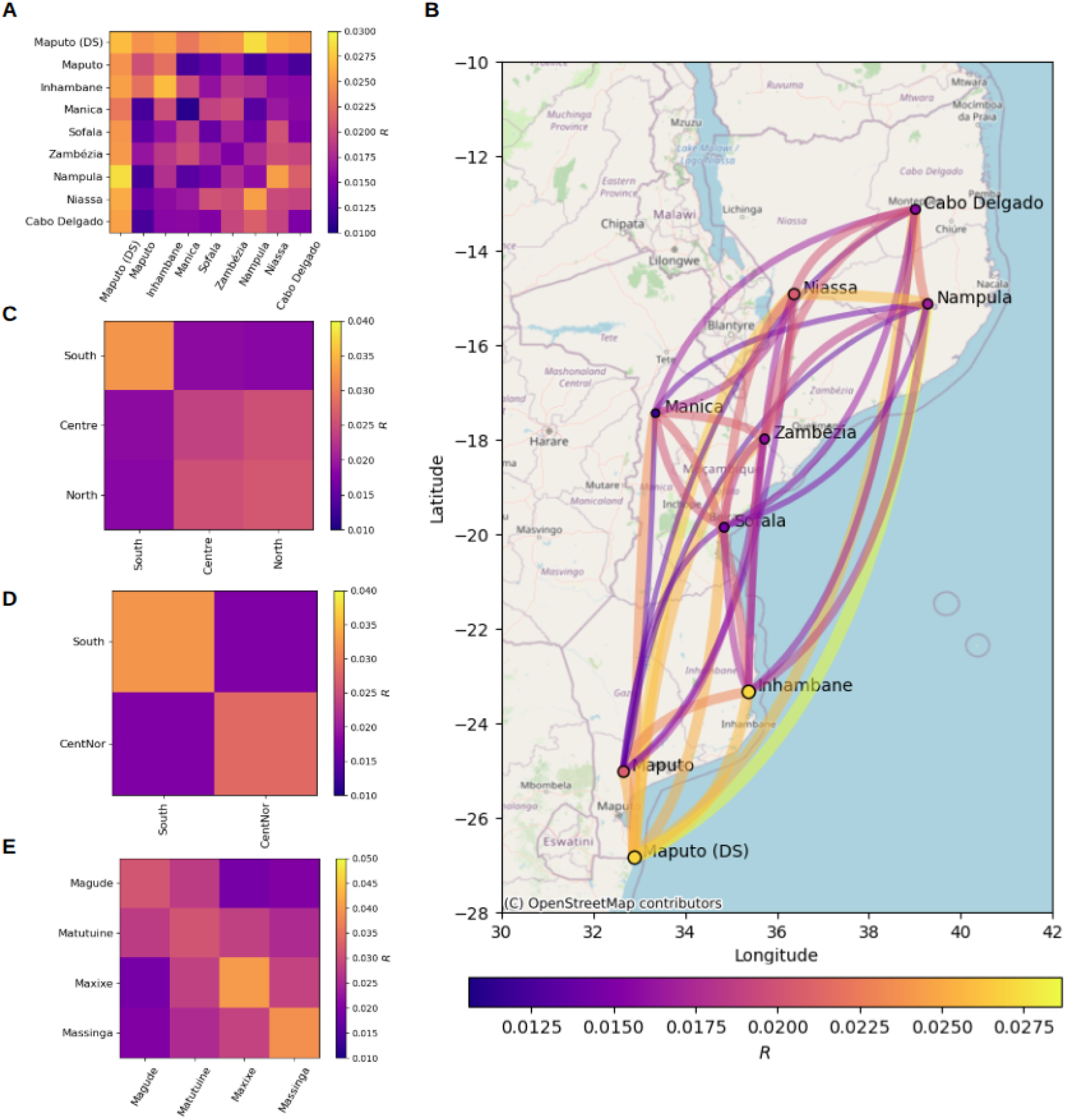
Genetic relatedness (identity-by-descent, IBD) of *P. falciparum* infections between regions in Mozambique. A) Fraction (*R*) of IBD-related sample pairs (IBD>0.1 with p<0.05) within and between different provinces, represented in colours. B) Spatial genetic connectivity between provinces. Line widths and point sizes are proportional to *R*, and colours show the ranking in *R* values (from blue to red, using turbo colormap). C) *R* between and within different regions (south, centre and north), combining samples from centre and north (D) and across Magude and Matutuine districts (E). N=1467. Maps used OpenStreetMap data, available under the Open Database License.

**Figure. 5:**
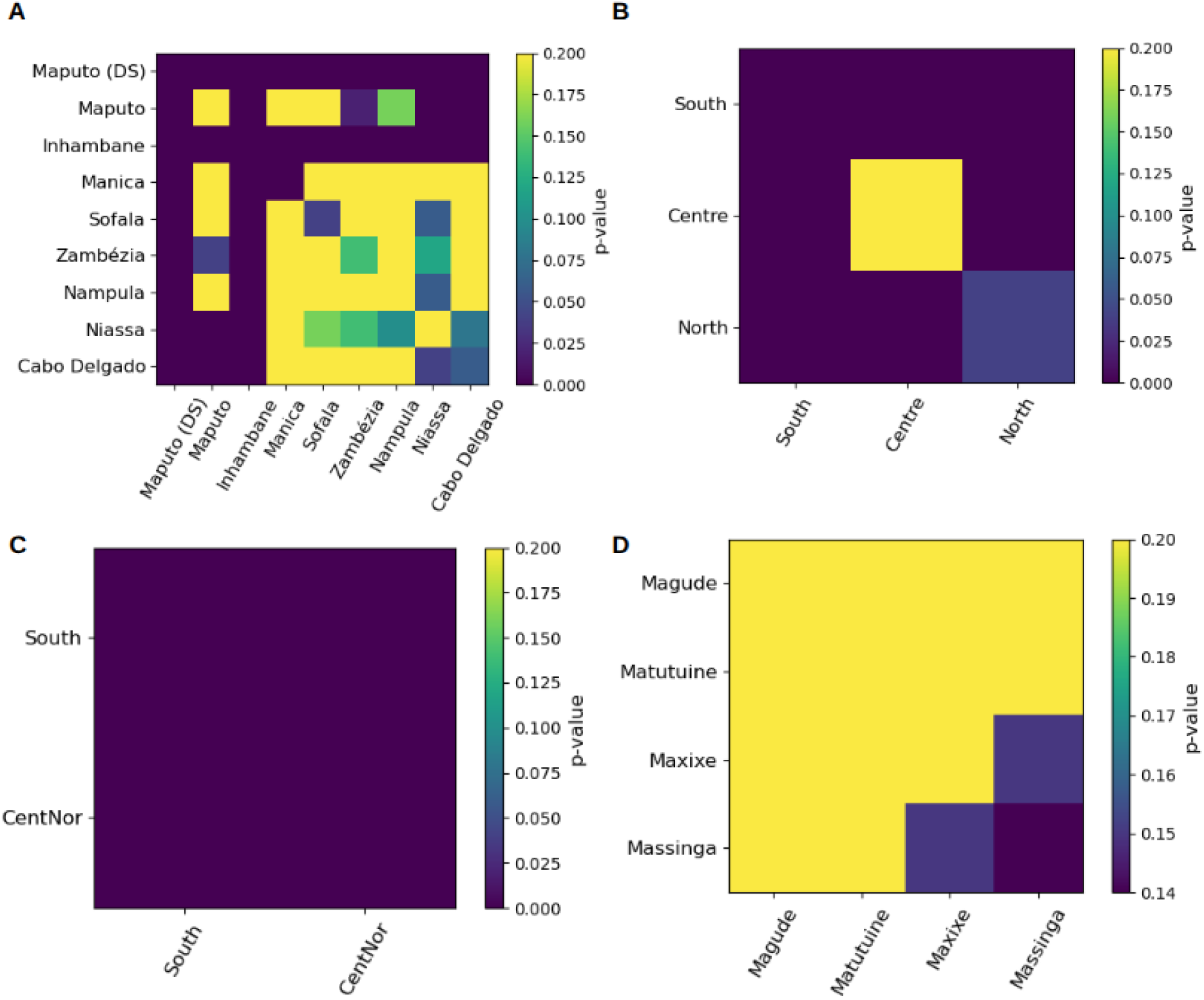
p-values of IBD-relatedness results. **A)** P-values of the deviation of the fraction of IBD-related pairs (IBD>0.1 with p<0.05) of the samples within and between different provinces with respect to the average across all pairs. **B)** The same, but comparing sample pairs within and across regions (south, centre and north). **C)** The same, but comparing sample pairs across south and centre/north. **D)** The same, but across Magude and Matutuine districts.

A strong spatial correlation of *R* at the inter-province scale was found (**Figure 6A**), with *R* significantly decreasing with the pairwise geographical distance for distances larger than 100 km (p<0.001). However, no significant correlation was found for distances between 10 km and 100 km (p=0.424, p=0.992 and p=0.454 when using IBD thresholds of 0.1, 0.15 and 0.2 respectively, showing that trends do not depend on the threshold used) (**Figure 6B**). In the shortest distances between zero (the same household) and 10 km, the decrease of *R* with distance became significant again (p<0.001 for IBD thresholds above 0.15) (**Figure 6C**).

**Figure 6:**
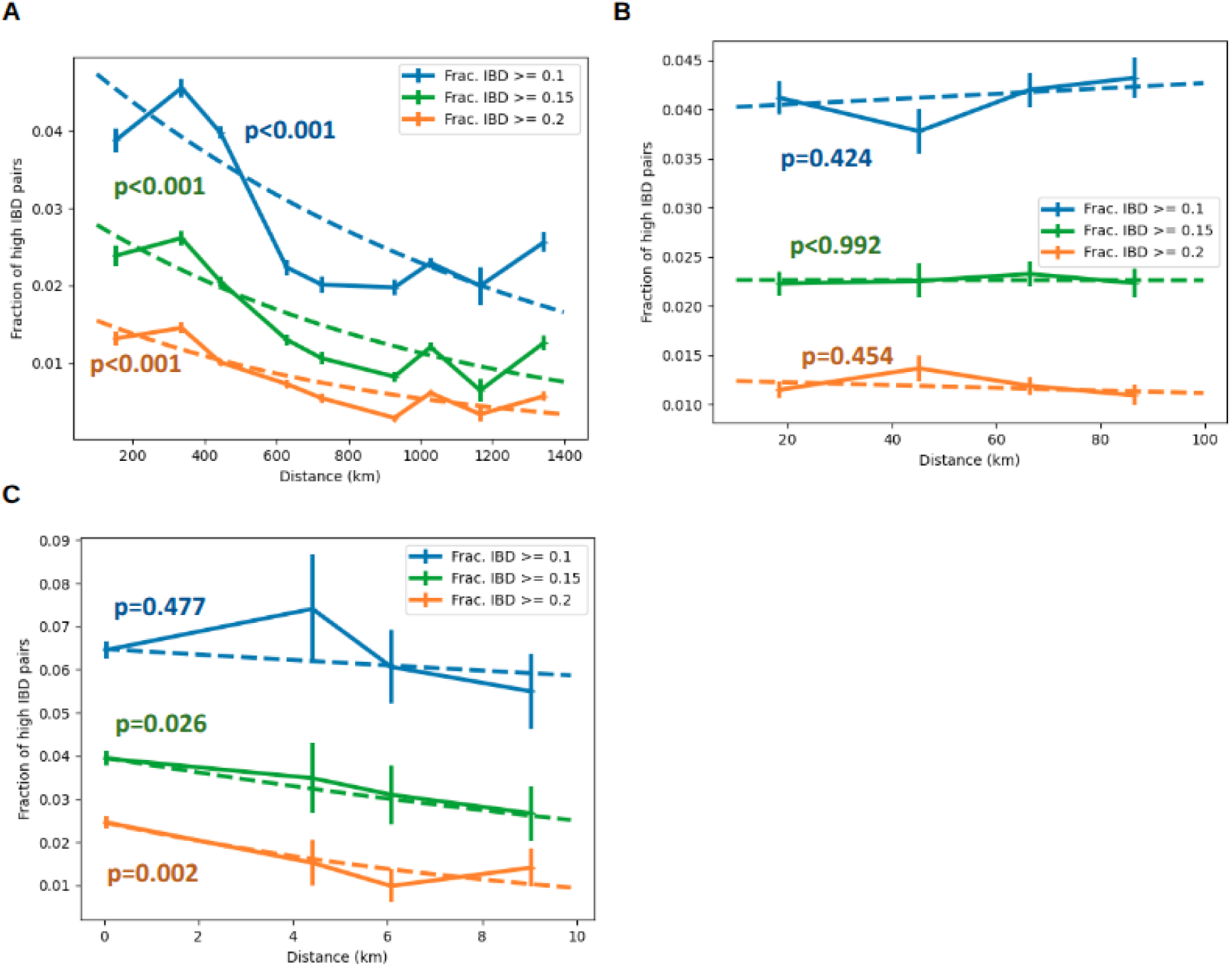
Genetic relatedness as a function of spatial distance. **A)** Fraction of IBD-related pairs (using different thresholds of IBD (always with p<0.05), shown in different colours as a function of the pairwise geographical distance, using a range of distances from 100 km to 1,400 km (typically inter-province samples). **B)** The same as **A** but for distances between 20 km and 100 km (typically within the same province but distinct health facilities). **C)** the same but for distances between 0 (same household) and 20 km, which is impacted by the significantly higher fraction for cases from the same household. N=1467.

### *P. falciparum* importation rates

A new Bayesian approach was used to classify clinical cases as imported and local. The method calculates an individual probability of being imported by combining epidemiological data (household-based mRDT positivity rates in children under 5 years per province from the demographic health survey 2022-2023^42^), travel reports (date, duration and destination, interpreting infections as local if no travels were reported) and genetic IBD relatedness (*R’* between the sample and the parasite population in Maputo province, or the travel destination, see Methods) (**Figure 7**). The importation probabilities obtained were in general close to 0 or 1, with only 1.5% (3/200) of them being between 0.25 and 0.75. Cases were classified as imported if their importation probability was higher than 50%. The fraction of imported cases within those who reported travel was 82.9% (87/105), corresponding to an importation rate of 43.5% (87/200) with respect to all cases. Among clinical cases from Magude and Matutuine with travel records, approximately half (54.3%; 57/105) reported a trip to Inhambane province, representing 28.5% (57/200) of all studied cases from Magude and Matutuine (**Table 4**). Similar results were found when travel duration (or *R’*) were not included in the estimation, being mRDT positivity rates the main driver of importation probabilities (**Appendix 1**).

**Figure 7:**
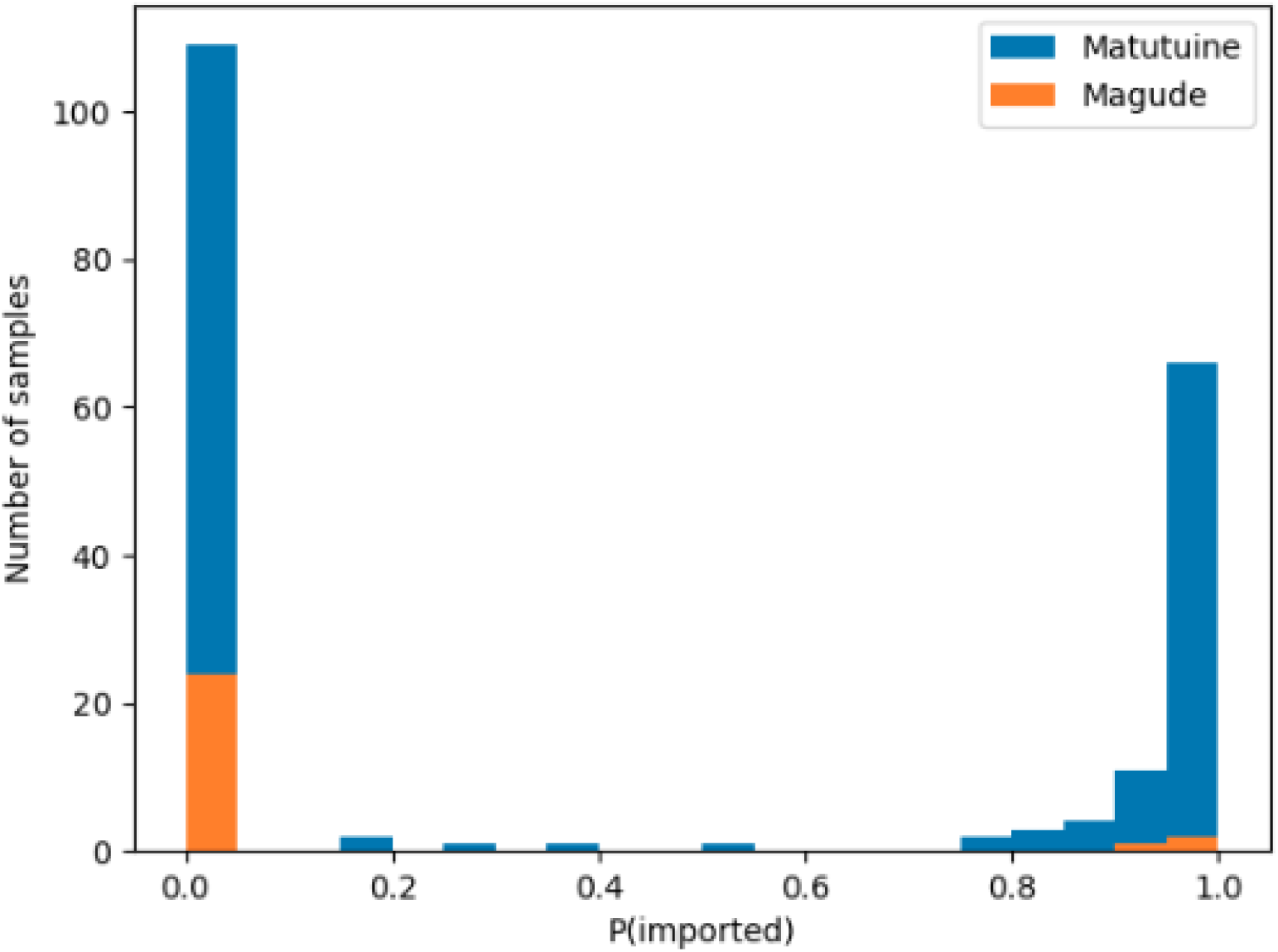
Distribution of importation probabilities by district. Distribution of the individual probabilities of being imported for the studied clinical cases from Magude (orange) and Matutuine (blue) districts.

**Table 4:**
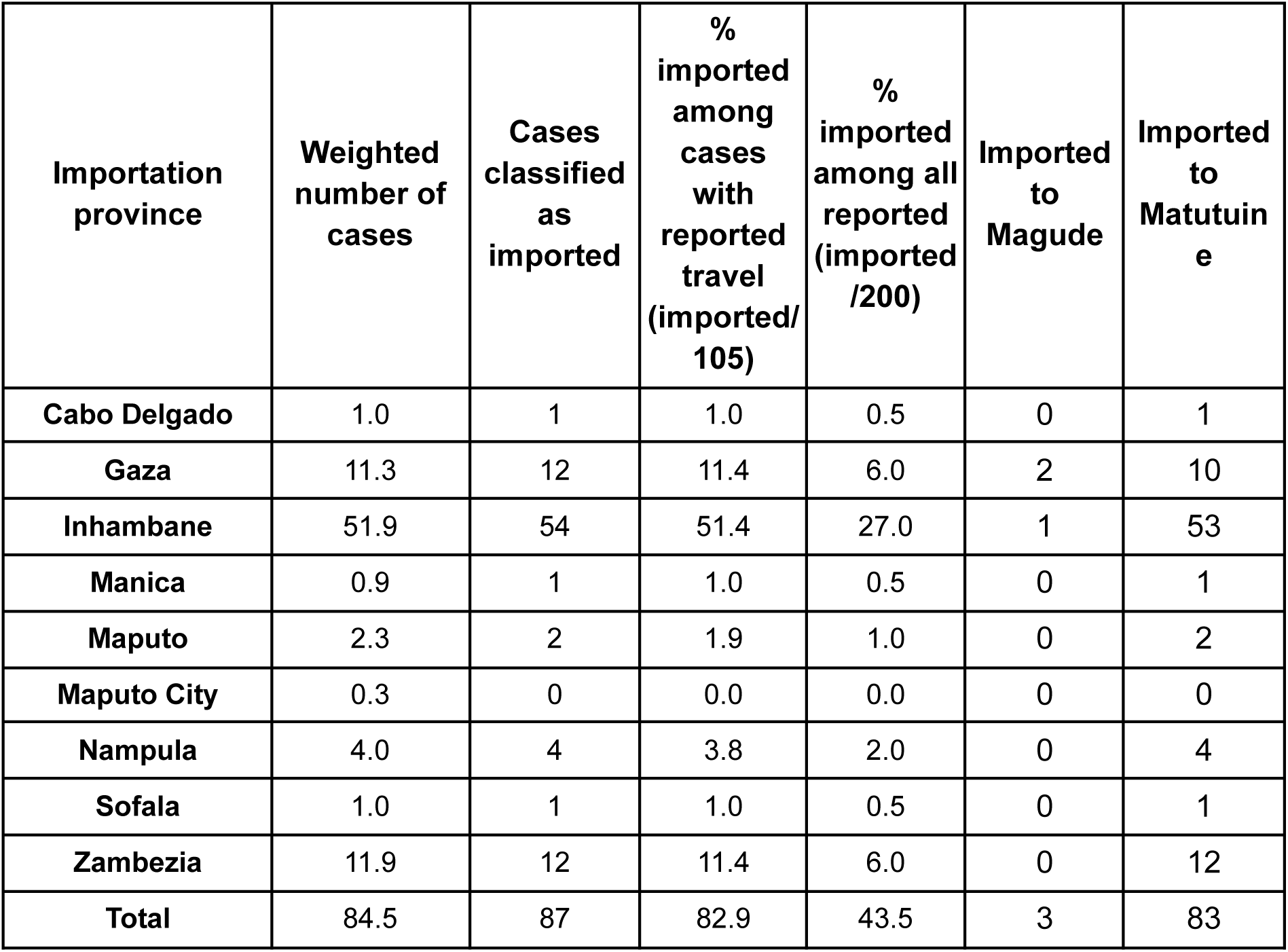
Travel destination (by province) of the imported cases reported in Magude and Matutuine. *Weighted number of cases*: the total number of cases (weighted by their probability) imported from each province (n=200). *Imported cases*: the total number of cases classified as imported if P(imported)>50% (n=200). *% cases with travel reports*: The fraction of imported cases from each province with respect to the total number of cases reporting travel in the previous 28 days (n=105). *% all reported*: the total contribution of imported cases from each province with respect to all reported cases in the Maputo province (n=200). *Imported to Magude*: total of imported cases residing in Magude (n=27). *Imported to Matutuine*: total of imported cases residing in Matutuine (n=173).

A statistically significant correlation was found between genetic relatedness and travel destinations. Cases reporting travel to Inhambane province presented a higher *R’* with the population from that province than cases reporting travels to other provinces or no travels at all (p=0.018) (**Appendix 2-Table 1**). This correlation was not statistically significant for other travel destinations, possibly due to the lower sample sizes of travellers (n<=15).

### Risk factor analysis of *P. falciparum* importation

Odds ratios (OR) were calculated using univariate and multivariate Firth’s logistic regressions^43^ to assess the risk factors associated with a malaria case being classified as imported (n=200; **Appendix 2-Table 2**, **Figure 8**). Pregnancy was excluded in the multivariate analysis due to the low number of pregnant women (n=2). The residence district was strongly associated with importation in both univariate (p<0.001) and multivariate (p=0.005) analysis, with Magude district presenting a lower proportion of cases classified as imported (11.1%, 3/27) than Matutuine (48.6%, 84/173; OR=6.6, 95%CI (2.3, 25.4)). Given that 96.6% (84/87) of the imported cases were from Matutuine district and only 3 were from Magude (**Table 4**), the statistics on imported cases, such as importation origin, mainly refer to Matutuine district.

**Figure 8:**
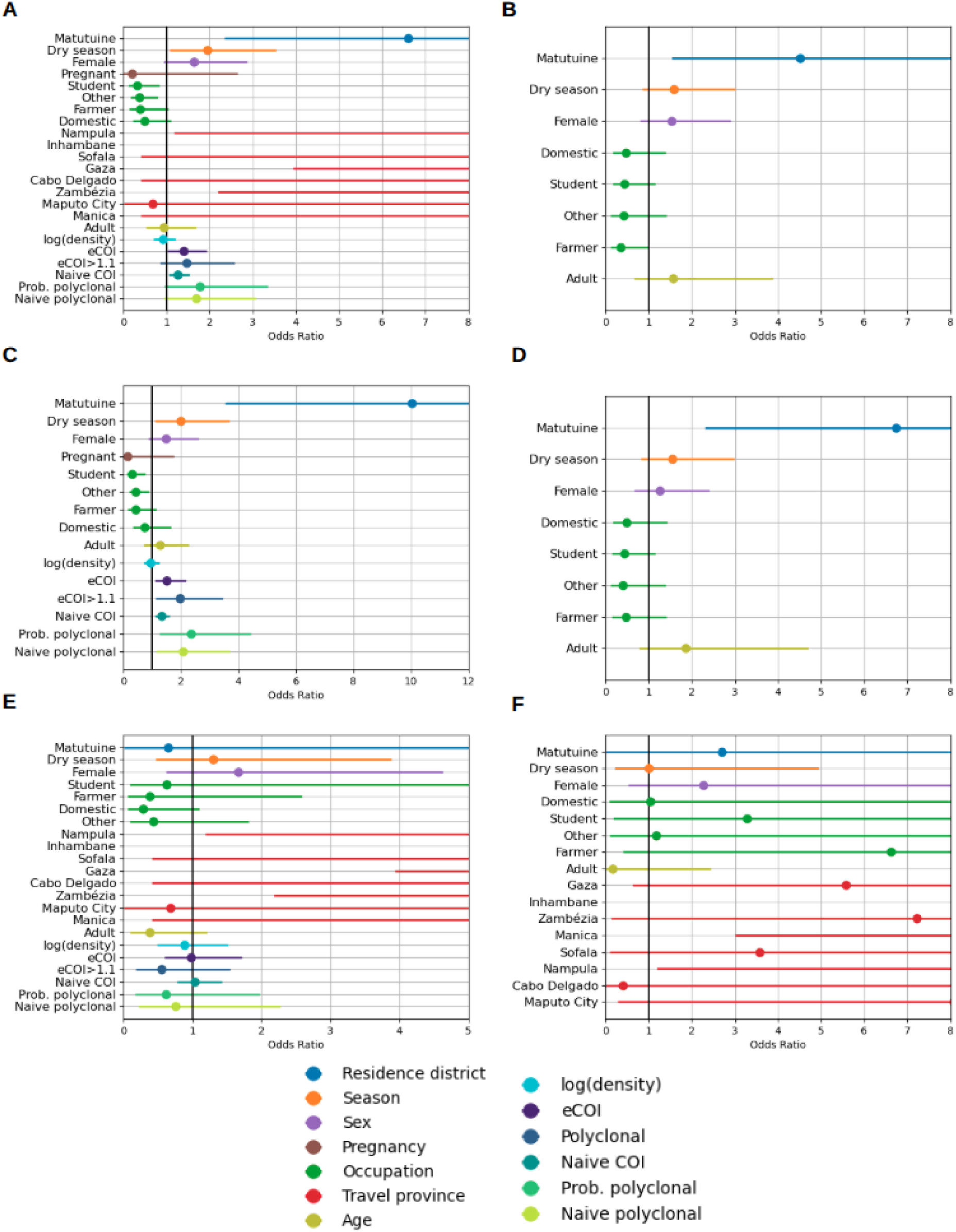
Odds ratio statistics of factors associated with importation and travel. Odds ratio of importation (A-B), reporting travel (C-D) and importation for cases with travel reports (E-F) for different factors in univariate (A, C and E) and multivariate (B, D and F) models, for all *P. falciparum* clinical cases recruited in Magude and Matutuine (n=200 for A-D, n=105 for E and F).

In the univariate analysis, season, occupation and travel destination were significantly associated with imported malaria. Cases occurring in the dry season had higher odds of being classified as imported compared to those in the rainy season (OR=1.9, 95%CI (1.1, 3.5), p=0.029), cases in students (OR=0.3, 95%CI (0.1, 0.8), p=0.019) and other occupations (OR=0.4, 95%CI (0.2, 0.8), p=0.011) had lower odds of being imported compared to unemployed (or minors), and cases having travelled to Gaza, Inhambane, Zambézia and Nampula had higher odds of being imported (>80%, p<0.04) (**Appendix 2-Table 2**, **Figure 8A**) compared to those having travelled to Maputo province. No associations were found for sex (p=0.085), pregnancy (p=0.246) or age (p=837). In the multivariate analysis, only the residential district remained a significant factor (p=0.005). No association was found between the odds of importation and parasite density (p=0.529) or any estimation of polyclonality (p>0.07 for any estimate). However, imported cases had a higher COI mean (2.80, 95%CI [1.0,7.8]) and eCOI (1.80, 95%CI [1.0,4.2]) than local cases (2.21, 95%CI (1.0,5.2), OR=1.3 95%CI=[1.1, 1.5], p=0.009 and 1.53, 95%CI (1.0,3.4), OR=1.4, 95%CI=[1.0,1.9], p=0.038, respectively) (**Appendix 2-Table 2**, **Figure 8A**). Similar results were found when assessing association between these factors and reported travel within the previous 28 days (**Appendix 2-Table 3**, **Figure 8C,D**).

When restricting the analysis to cases reporting travel (n=105), the only factor that remained significantly associated with being classified as imported was the province destination of the travel (p<0.034 for the province destinations of Gaza, Inhambane, Zambézia and Nampula), from both univariate and multivariate analysis (**Appendix 2-Table 4**, **Figure 8E,F**). The probability of being imported was above 80% for all travels reported outside Maputo province and Maputo city, where the importation probability was below 20%. Neither parasitemia, COI nor polyclonality were found to be associated with importation.

## Discussion

Knowledge of the sources of transmission, including imported cases, can inform the tailoring of effective targeted approaches for elimination^40,44^. Here we have developed a novel Bayesian approach that allows the integration of genetic, travel history and epidemiological data to estimate probabilities of malaria importation. This new approach combines pairwise IBD estimates with travel reports to obtain a case-by-case probability of being imported. We found different importation rates between Magude (11.1%, 3/27) and Matutuine districts (48.6%, 84/173; OR=6.6, 95%CI (2.3, 25.4), p<0.001), in Maputo province, identifying Inhambane province as the main transmission source in Matutuine. COI and eCOI, which are within-host markers potentially informative of malaria burden in the population^45^, were higher for imported cases, confirming our importation estimates from areas of higher malaria burden.

The results show that parasite genomics can be used to assess genetic population structure at the national level and at very-small scales (at the household level), but not at district level. We observed a high genetic relatedness between *P. falciparum* sample pairs within the south and within the centre-north of the country and a lower relatedness between both regions. Pairwise genetic IBD-relatedness significantly decreased with the distance at large geographical scales, suggesting a strong isolation-by-distance at the country level, probably due to different parasite populations having different allele frequencies. No spatial correlation was found between samples at distances between 10 and 100 km, representing intra-province distances, indicating that *P. falciparum* genomics is heterogeneous at the provincial level. However, the spatial correlation was significant again at the smallest scales, including the distance of zero (the same household). This indicates that genetic IBD-relatedness is significantly higher for intra-household pairs, quickly decreasing to show no spatial pattern at small distances. The high relatedness of household members highlights the potential of genomics for very fine-scale transmission modelling approaches, such as transmission network modelling^46^. The large-scale genetic structure suggests that *P. falciparum* genomics can be used to assess malaria importation across provinces, but the lack of structure at distances of 10-100 km, maybe due to high mobility in short distances, might challenge the studies on importation within the province (across districts).

Cases notified during the dry season reported more travels (64.1%) and showed higher importation rates (54.7%) than those from the rainy season (47.1% and 38.2% respectively). However, there was no significant association with importation when conditioned to travel, implying that that importation and travel are highly correlated. This indicates that importation is higher precisely when transmission is lower. No association with importation was found for occupation, sex, age or pregnancy, suggesting that targeting specific subpopulations for malaria prevention or control might not be strategic.

A high fraction (52.5%) of *P. falciparum* clinical cases in Magude and Matutuine districts reported a travel in the last 28 days. Among them, a total of 82.9% of these cases were estimated as imported, representing 43.5% of all reported cases studied. Remarkable differences in importation rates were found between Magude (11.1%) and Matutuine (48.6%) districts, as well as in travel rates (11.1% in Magude and 59.0% in Inhambane). Inhambane was found to be the main source of importation in Matutuine, accounting for 63.9% (53/83) of all the imported cases from Matutuine. Genetic relatedness confirmed these mobility patterns, with Matutuine showing higher genetic relatedness with Inhambane than Magude. Several factors can contribute to these different importation patterns. Matutuine district is close to Maputo City (a city with high mobility and importation levels), has better communication infrastructures and has some touristic places (e.g. Ponta do Ouro) of potential importation risk. In contrast, Magude district is an interior rural area, more isolated from big communication hubs, with less mobility due to work and with the National Kruger Park (South Africa) limiting movement in a significant fraction of its border. Further social studies would be required to identify those factors that increase the importation risk in southern Mozambique.

The results of this study have several practical implications for malaria elimination. First, improving the case classification through the obtained probabilities of importation conditional to the reported travel destinations allows for a better quantification of importation in elimination districts. Second, preventing importation, either by testing and treating travelers or by reducing malaria transmission in Inhambane, might be especially relevant to eliminate malaria in Matutuine district. These efforts would be more cost-efficient during the dry season when importation rates are higher. These interventions could potentially target around 60% of the infections in Matutuine, given that 65.5% of the studied cases are genetically more related to Inhambane than to Maputo. However, a better understanding of transmission networks is needed to quantify the real impact of importation on the overall transmission. Finally, targeting the whole population through vector control or mass drug administrations (MDAs) may be more appropriate in areas such as Magude where importation rates are low. In both districts, reactive strategies targeting remaining infections will be needed to interrupt transmission^47^.

While these results propose programmatic strategies for the two study districts, routine surveillance to detect importation in Mozambique would allow for identifying new strategies in other districts aiming for elimination, as well as monitoring changes in importation rates in Magude and Matutuine in the future. If scaling molecular surveillance is not feasible, travel reports could be integrated in the routing surveillance to extrapolate the case classification based on the results of this study. In general, the method presented here can be applied to other *P. falciparum* endemic areas, as well as to *P. vivax* endemic areas outside Africa. For the case of *P. vivax*, symptomatic cases are not necessarily reflecting recent infections, so that travel reports might need to cover longer time periods, which does not require any essential adaptation to the method.

The study presents some limitations. First, except in Magude and Matutuine, *P. falciparum* isolates in the rest of the country were collected at selected health facilities and therefore may not be representative of the whole parasite population circulating in Mozambique, potentially biasing genetic relatedness with respect to Maputo province and potentially overestimating the genetic differentiation and underestimating importation. In particular, in Nampula, the sampling was conducted within 2 weeks in January and at only one health facility per district. Also, assuming that the samples from each province are representative of the travel destinations of the study cases might underestimate importation. However, the impact on our results must be small due to the high importation rates obtained. Second, only 24.7% (200/809) of the index cases were included in the importation analysis due to the low completeness of metadata, which might also induce some unknown biases. Third, our proxy of malaria transmission intensity was based on mRDT positivity rates in children under 5 years from a household-based survey, which is optimal for estimating malaria burden but not for transmission intensity. Better proxies of transmission intensity could be malaria incidence at the monthly level from national surveillance systems, or estimates of force of infection, for example from the use of molecular longitudinal data if available. Also, importation probabilities relied on travel reports and were sensitive to their potential biases (e.g. unreported travels). Finally, seasonality of malaria transmission was not taken into account in the modelling of importation probabilities, which could increase the precision of the estimates. Future work to address some of the limitations includes: studies increasing sample sizes with data from 2023 to 2025 with a cluster sampling approach to address sample representativity; and refining a model of monthly malaria incidence.

To conclude, a new Bayesian model was used combining epidemiological, mobility and genetic data simultaneously to provide individual case classification, which potentially allow for identifying individual factors and specific populations for fine targeted approaches for elimination. Both mobility and genetic data were found to be informative of importation, highlighting the potential of malaria genomics to refine importation estimates. Very distinct importation and travel rates were found in two close districts with very similar malaria burden. The main sources of transmission were identified in these low-transmission areas that can inform decision-making strategies for malaria elimination.

## Methods

### Study design and sample collection

In Magude and Matutuine districts, all clinical cases who tested positive for malaria using HRP2-based rapid diagnostic tests (Bioline Malaria Ag *P.f.*, 05FK50 [Abbott], First Response® Malaria Antigen *P. falciparum* HRP2 [Premier Medical], AdvDxTM Malaria *Pf* Rapid Malaria Ag Detection Test [Advy]) during 2022 were invited to participate in the study if they were older than 6 months, resided in the area and had no symptoms of severe malaria. For all participants giving consent (by their adult representative for minors), forms were filled out to collect demographic and epidemiological information, including travel within the past 28 days (with dates and destinations). The RDTs were labeled with patient ID codes, placed in zip-lock plastic bags containing silica gel, and shipped to CISM where they were stored at –20°C until further processing.

In addition, a convenience sampling approach was conducted at selected health facilities in 9 provinces during the rainy season (January to May) of 2022^38,40^: Maputo (including Maputo city; all ages >6 months); Inhambane, Manica, Zambézia, Sofala and Manica (children 2-10 years old); Nampula (children 3 months-5 years old); and Tete and Cabo Delgado (children 6 months-5 years old) (**Figure 1**). Inclusion criteria were confirmed diagnosis of uncomplicated malaria by routine RDT, providing informed consent (participant or adult representative). For Tete and Cabo Delgado samples were collected at baseline as part of antimalarial therapeutic efficacy study 2022, with an additional inclusion criteria of >1000 parasites/μL by light microscopy. Once consent was given, two to four 50μL dried blood spots (DBS) were prepared onto one or two filter papers through finger prick. DBS were identified with anonymous barcodes, air-dried, packed with silica gel and stored at 4°C until shipment to CISM, where sealed bags were kept at –20°C in the central laboratory until processing. The number of successfully sequenced samples per province ranged from 44 to 364, being the highest number for Inhambane province, the most frequent travel destination of the study cases (**Table 3**). All samples were shipped to ISGlobal (Barcelona) for sample processing.

### Genomic DNA extraction and quantification

Genomic DNA was extracted from RDT strips or from DBS samples using a Tween-Chelex based protocol^39^. To extract DNA from RDTs, cassettes were open and the nitrocellulose strip was separated using sterile forceps. Both the conjugate pad and the proximal part of the nitrocellulose strip were cut using sterile scissors in small pieces and placed into a well of 96-deep well plates. In the case of DBS, 5 mm (∼12,5 μl blood) discs were cut from each DBS with a manual puncher and placed into 96-well deep well plates. One ml of freshly made 0.5% Tween 20® detergent diluted in PBS was added to each well plate containing a DBS punch and incubated overnight in a thermomixer at 15°C and 300rpm. The next morning, the supernatant was removed, 1 ml of fresh PBS was added per well and the plate was briefly vortexed and then incubated at 4°C for 30 min. After incubation, the liquid was aspirated, and 150 μl of a solution of 10% Chelex (C7901, Merck) in molecular grade water. The samples were incubated at 95°C in a water bath for 15 min with gentle vortexing every 5 min. The plate was then centrifuged for 5 min at 1500 rpm to pellet the Chelex® beads. Supernatant (approximately 130 ul) containing the eluted gDNA was transferred to a new PCR 96 well plate, centrifuged again and finally 100 ul transferred to barcoded plates (Wilmut). Positive (3D7, MRA-151; or HB3, MRA-155; or Dd2, MRA-156; or Dd2_R539T, MRA-1255 [MR4, Bei Resources]) and negative (non-infected blood) controls prepared as RDT or DBS were added to each plate, depending on the type of sample being processed.

*P. falciparum* infection was confirmed in all DNA samples by qPCR targeting the 18S rRNA gene on an ABI PRISM 7500 HT Real-Time System (Applied Biosystems), as previously described^48^. Parasite density was quantified by extrapolation to an external standard curve composed of six 1:10 dilutions of 3D7 cultured parasites in whole blood (range 1 to 100,000 parasites/µl; MRA-151, MR4, Bei Resources). DNA was stored at –20°C until sequencing.

### Amplicon-based sequencing and sequence data analysis

Sequencing was performed using the MAD^4^HatTeR multiplex amplicon sequencing panel as previously described^38,39^. Multiplexed PCR primer pools D1, R1.2 and R2 were used, which combined target 241 *P*. *falciparum* loci of 225-300 bp^41^. DNA was amplified in a multiplexed PCR (Paragon Genomics Inc, California, USA) for 10 (if parasite density ≥500 parasites/µL) or 20 cycles (<500 parasites/µL). A randomly selected subset of 8 libraries from each full 96-well plate was assessed using automated electrophoresis in a TapeStation 4150 (Agilent Technologies, California, USA) to confirm library quality and specificity of products. Libraries were pooled adjusting volumes based on parasitemia, bead-cleaned using CleanMag® Magnetic Beads (Paragon Genomics Inc, California, USA) to remove primer dimers and run on an agarose gel, from which the amplicon-sized band was excised (Monarch® DNA Gel Extraction Kit, New England Biolabs Inc., Massachusetts, USA) and quantified with Qubit™ 1X dsDNA High Sensitivity assay kits. Pools of 288 samples were sequenced with 150 paired-end reads in a NextSeq 2000 System using P1 reagents (Illumina, USA). Positive (DBS prepared with *P. falciparum* laboratory strains 3D7 (MRA-151), HB3 (MRA-155), Dd2 (MRA-156 and MRA-1255)) and negative controls were included in every library preparation plate.

FASTQ files were subjected to filtering, demultiplexing and allele inference using MAD^4^HatTeR Nextflow-based pipeline version 0.1.8 (https://github.com/EPPIcenter/mad4hatter)^41^. The 3D7 genome sequence was used as reference for alternative allele calling (https://github.com/EPPIcenter/mad4hatter/blob/main/resources/v4/ALL_refseq.fa). The resulting allele tables were subsequently filtered based on read counts and coverage across loci within a sample and across samples. Alleles with fewer reads than the maximum observed reads in any locus for negative controls were removed, along with alleles with <1% within-sample frequency. To identify potential issues of duplications in sample collection and library preparation, sample similarity was estimated using the root mean square error of within-sample allele frequency (WSAF) for highly diverse targets (pool D1.1), followed by clustering by Density-Based Spatial Clustering of Applications with Noise (DBSCAN) with ε=0.2. Clusters of samples (at least two samples) with clonality >2 and nearly identical WSAF across all loci, suggesting multiple DBSs were prepared using blood from a single patient, were excluded from the analysis.

For this analysis, the 165 loci of diversity from MAD^4^HatTeR D1 pool were used. Samples with less than 75% loci (n=123) covered with a minimum of 100 reads were excluded. The exclusion criteria for loci was defined as those with less than 100 samples covering at least 100 reads.

Intra-host COI, eCOI and polyclonal probability were obtained using MOIRE v3.4.0^49^ (https://github.com/EPPIcenter/moire), which uses a Monte Carlo Markov Chain (MCMC) approach taking into account intra-host relatedness and allele frequencies at the population level. MOIRE was run taking all samples as one populations, but no significant differences were found when run across different regions. Naive polyclonality was defined as COI>1, and polyclonality as eCOI>1.1. Genetic diversity was estimated as IBD using Dcifer 1.2.0^50^ (https://eppicenter.github.io/dcifer/), which takes into account polyclonal infections and their intra-host relatedness to infer IBD across sample pairs. Dcifer provides an estimate of IBD as well as a p-value of rejecting IBD>0. In the analysis, sample pairs were defined as IBD-related if IBD>0.1 (higher thresholds implied lower statistics of related pairs) with p<0.05 (rejecting IBD=0), and the genetic relatedness, *R*, between two populations (or within a population) was defined as the fraction of IBD-related pairs between (or within) populations, using only the compared populations to estimate *R* (equivalent trends were found for different IBD thresholds).

### Statistical analysis

IBD analysis was conducted across provinces and across regions, defining regions as south (Maputo, Inhambane and Gaza), centre (Manica, Sofala, Tete and Zambézia) and north (Nampula, Niassa and Cabo Delgado). The statistical significance of *R* between (and within) populations was calculated as their difference with respect to the average *R* across the whole population. The p-value was calculated from bootstrap resampling with replacement, assuming no spatial differentiation.

Spatial structure of *R* (using IBD thresholds of 0.1, 0.15 and 0.2) was studied as a function of the pairwise geographical distance, using different distance ranges (100-1,400 km, 10-100 km and 0-10 km), with binning balancing spatial granularity with sample size. The statistical trends were obtained from a logistic regression of *R* versus distance, with a p-value of rejecting no dependence.

A new Bayesian approach was used to infer probabilities of importation combining epidemiological, genetic and mobility data. The approach models the probability of being infected (*I*) for *P. falciparum* in an area (*A*) given the *P. falciparum* genome (*G*) of the infection as:

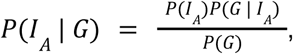

Where *P*(*I_A_*| *G*) is the probability of being infected in area *A* given a *P. falciparum* genome *G*, *P*(*G* | *I_A_*) is the probability of having a *P. falciparum* genome *G* if the infection occurred in area *A*, and *P*(*I_A_*) and *P*(*G*) are the probabilities of being infected in area *A* and having *P. falciparum* genome *G* respectively. To estimate *P*(*I_A_*), it was assumed that the probability of an infection to have occurred in a province is proportional to a) the time spent in that province, and b) the transmission intensity of that province:

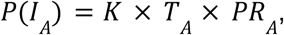

Where *T_A_* is the time spent in area *A*, *PR_A_* is (a proxy of) the transmission intensity in *A* and *K* is an unknown constant that does not depend on the specific area, assuming that the differences in transmission intensity were captured in *PR* (**Appendix 1**). *P*(*G* | *I_A_*) was estimated as *R’_A_*, defined as the fraction of samples from *A* that are IBD-related to the sample studied. With the constraint that *P*(*I_A_*| *G*) + *P*(*I_B_*| *G*) = 1 if the case had only stayed in two areas recently (as the study cases), *P*(*I_A_*) becomes:

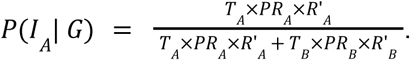

The probability of being imported was obtained defining *A* as the travel destination area and *B* as the local area (Maputo province). Assuming that the infection did not occur earlier than the past 28 days, a case was considered locally transmitted if it reported no travels in the last 28 days. The infection was assumed to take at least 7 days to become symptomatic, so *T_A_* and *T_B_*were obtained from the last 7 to 28 days. For missing travel durations, the average *T* of all available data was imputed, corresponding to 9.14 days. *PR_A,B_* were estimated from the mRDT positivity rates (PR_RDT_) per province in children reported in the last Health Demographic Survey 2022-2023 from Mozambique^42^. Since the PR_RDT_ in Maputo City was rounded as 0.0, a value of 0.04 was assumed, probably overestimating (with negligible impact) the infection probabilities in Maputo City. *R’_A,B_* was estimated as the fraction of samples from the area (province) that were IBD-related (IBD>0.1, p<0.05) with the sample studied. When estimating *R’* with Maputo province, one could consider excluding the samples from clinical cases reporting trips to avoid biases from imported cases. The impact of this choice was lower than 1% in the estimated imported rates, so all cases were included to estimate *R’*, a more conservative approach that was also more consistent with the other provinces where no travel reports were available. For missing *R’* estimates, such as in Gaza province, only *T* and *PR* factors were used to estimate probabilities of importation. Cases were classified as imported for importation probabilities higher than 50%, and as local otherwise. Importation rates were obtained from the fraction of imported cases over all classified cases. Similar rates were obtained using weighted sums of individual probabilities given the extreme values of probabilities obtained (**Table 4**). In case of obtaining a higher fraction of intermediate values (0.4-0.6), weighted sums of individual probabilities would be more appropriate to better quantify importation rates.

The correlation between travel reports and genetic relatedness was quantified from the fraction of cases reporting (or not) a travel in a given province and their fraction of cases that were more related to the origin and travel destination populations, conducting a chi square test for independence under the assumption that the probabilities of being more related to an area than to Maputo province were not correlated with travels reported.

Risk-factor analyses were conducted using firth logistic regression to identify factors associated with importation and travel. The analysis included the following characteristics: district of residence (Magude or Matutuine), seasonality (defined as rainy for cases reported from January to May and dry for cases from June to December), sex, pregnancy, occupation, age (stratified as adults and minors) and province of travel destination. In order to assess the potential of molecular data to inform about imported cases, the same analysis was done including parasite density, eCOI and polyclonality. Since imported cases required a travel report by definition, the same factor analysis on importation was conducted restricted to those cases with travel reports (this time including travel destination in the multivariate analysis). Odds ratios and p-values were obtained from the firth logistic regressions in a univariate analysis and also in a multivariate analysis using all factors without interactions (the limited sample sizes did not allow for exploring interactions between factors).

All over the analysis, statistical significance was defined as p<0.05. All the analyses were performed using Python 3.9.16, Jupyter Lab 3.3.2 and R 4.2.3.

### Ethical considerations

This study was conducted in accordance with the ethical principles outlined in the Declaration of Helsinki. Ethical approval was obtained from the Comité Nacional de Bioética para Saúde (CNBS) Mozambique, affiliated with the Ministério de Saúde (approval number 604/CNBS/21; 1st November 2023). The CNBS can be contacted at: Ministério de Saúde – 2° andar dto, Av. Eduardo Mondlane / Salvador Allende, PC 264, Maputo, Mozambique.

## Data availability

Sequencing data is available at NCBI Sequence Read Archive (SRA) under the accession numbers specified in the **supplementary file 1**.

## Code availability

All code used in the analysis is open source under a GNU General Public License. The main open source repository of this analysis can be accessed here: https://github.com/MalPhyGen/malaria_relatedness_importation. The repository contains software requirements and installation instructions, with references to other public repositories used, and demos of the whole analyses conducted.

## Supporting information

Sample list

## Acknowledgments

We would like to thank all the individuals of the study and their corresponding parents/guardians who agreed to participate in the study, clinical officers, field supervisors, data managers and lab technicians from all participating institutions. We thank Llorenç Quintó for his advice on statistical analysis. This work was supported financially by the Bill and Melinda Gates Foundation (INV-019032, A.M. and INV-067310, A.M.), the Government of Catalonia and co-financed by the European Social Fund of the EU (Agència de Gestió d’Ajuts Universitaris i de Recerca [AGAUR] grant 2022 FI_B 00148, S.B. and SGR 01517 to A.M.), the European Union’s Horizon 2020 research and innovation programme (Marie Skłodowska-Curie grant 890477, A.P.). The project that gave rise to these results received the support of a fellowship from the “laCaixa” Foundation (ID 100010434). The fellowship code is LCF/BQ/PR24/12050009 (A.P.). The Centro de Investigação em Saúde de Manhiça (CISM) is supported by the Government of Mozambique and the Spanish Agency for International Development Cooperation (AECID). This research is part of ISGlobal’s Program on the Molecular Mechanisms of Malaria which is partially supported by the Fundación Ramón Areces. We acknowledge support from the grant CEX2023-0001290-S funded by MCIN/AEI/10.13039/501100011033, and from the Generalitat de Catalunya through the CERCA Program. The funders of the study had no role in study design, data collection, data analysis, data interpretation, or writing of the report.

## Competing interests

The authors declare no competing interests.

## Appendix 1 Supplementary importation analysis

In very-low transmission settings, identifying the key sources of transmission can be crucial for effective elimination efforts. Importation can contribute to sustain transmission in these areas from two transmission steps. First, the introduction of individuals in the area that were previously infected elsewhere. These are infected cases that would ideally be classified as imported. Second, the local spread of this imported parasite in the community, which will increase local transmission with the newly introduced parasite. When studying malaria importation, one might want to conduct case classification in order to identify imported and local cases, or to quantify the impact of importation on the overall transmission, in which case all infections from the two steps described should be included.

Both travel history data and parasite genetic relatedness provide information about malaria importation and its impact, but each data source comes with different limitations. In the case of travel reports, they have the potential to identify the possible origins of malaria infections, but biases in their reporting (unmentioned trips, lack of data from older than 28 days ago, etc.) have a direct impact on the considered possibilities. On the other hand, parasite genetics have the potential to assess the origin of the parasite without any need of mobility data, avoiding their biases, but would not allow for distinguishing between imported cases and local spread after imported case.

The aim of this study was to conduct case classification, leaving the quantification of its impact on the overall transmission for a future study. To this aim, the formula described in the main manuscript was used, according to the model described:

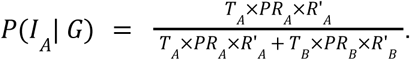

This model relies on travel reports to define the possible origins of infections, so that importation is only contemplated to happen during the travels reported in our study. It also uses genetic relatedness metrics to refine the probabilities of infections in the different areas. This approach comes with the limitations mentioned above regarding biases on travel reports, but avoids estimating origins of infections (from genetics) from areas with no evidence of travels. In this sense, the method can be interpreted as a conservative case classification method that integrates parasite genomics, travel history data and epidemiological data simultaneously.

The specific application of this model comes with several assumptions and limitations regarding each of the metrics used. First, *T_A,B_* was defined as the time spent in areas *A,B* between 7-28 days ago. This assumes that infections only happened within this period, and that the probability of infection is constant over this time. This estimation could be refined from modelled probabilities of infection age for symptomatic cases, and improved with travel reports including older periods. Second, *PR_A,B_* were estimated from the mRDT positivity rates (PR) per province in children reported in the last Health Demographic Survey 2022-2023 from Mozambique^1^. This assumes that PRs are a robust proxy of the transmission intensity (or force of infection), and that they are linearly related, which might not hold especially in extreme cases^2^. The model would benefit from better estimates of transmission intensity, considering also finer time granularities, such as monthly estimates of incidence. Finally, the model assumes that the probability of having a genetic data *G* when being infected in area *A* is given by *R’_A_*, so the genetic relatedness (fraction of genetically related pairs) between the case and the parasite genetic population from *A*. If the parasite genetic population in *A* is well represented, an random infection from *A* would have equal chances to be of any of its parasites. *R’_A_* is taken as a proxy of the probability of being any of them, which is not strictly true, but it is a feasible approximation to the ideal scenario, which would be calculating the fraction of clonal pairs between the study case and the whole parasite population in *A*. The model used assumes that *R’_A_* is linearly proportional to this ideal estimate. Future analyses with simulated data would be useful to confirm this hypothesis. Finally, modelling the robustness of travel reports would allow for including probabilities of other sources of transmission than just those reported.

In order to analyse the contribution of each data source (*T_A,B_*, *R’_A,B_* and *PR_A,B_*) on the final probabilities of importation, these were calculated using different modifications of the model: using the original formula; ignoring the information on travel duration (forcing *T_A_*=*T_B_*); ignoring the information on genetic relatedness (forcing *R’_A_*=*R’_B_*); ignoring the epidemiological data (forcing *PR_A_*=*PR_B_*); and for combinations of them, ignoring two factors simultaneously. **Appendix 1-Figure 1** shows the distribution of probabilities of importation (for mRDT+ cases with travel reports) for the original model (black) and the models without the factors *R’_A,B_* (orange), *T_A,B_* (purple) and *PR_A,B_* (green). **Appendix 1-Table 1** describes the overall importation rates from the weighted sum of probabilities of importation for the different approaches. *PR_A,B_* was found to be the main driver of case classification, with the majority of mRDT+ cases showing importation probabilities close to one when *PR_A,B_* was included in the estimator. This reflects the high differences in PR between the local province and most of the travel destination provinces. Only when *PR_A,B_* was excluded the importation probabilities generally decreased, with very few cases showing probabilities of one. The inclusion of *T_A,B_* and *R’_A,B_*contributed to slightly reduce the importation rates (to <90% of cases with travel reports) with respect to using only *PR_A,B_* estimates (91.786% of cases with travel reports), suggesting that *T_A,B_* and *R’_A,B_* contributed to refine the estimates.

---

**Appendix 1-Figure 1:**
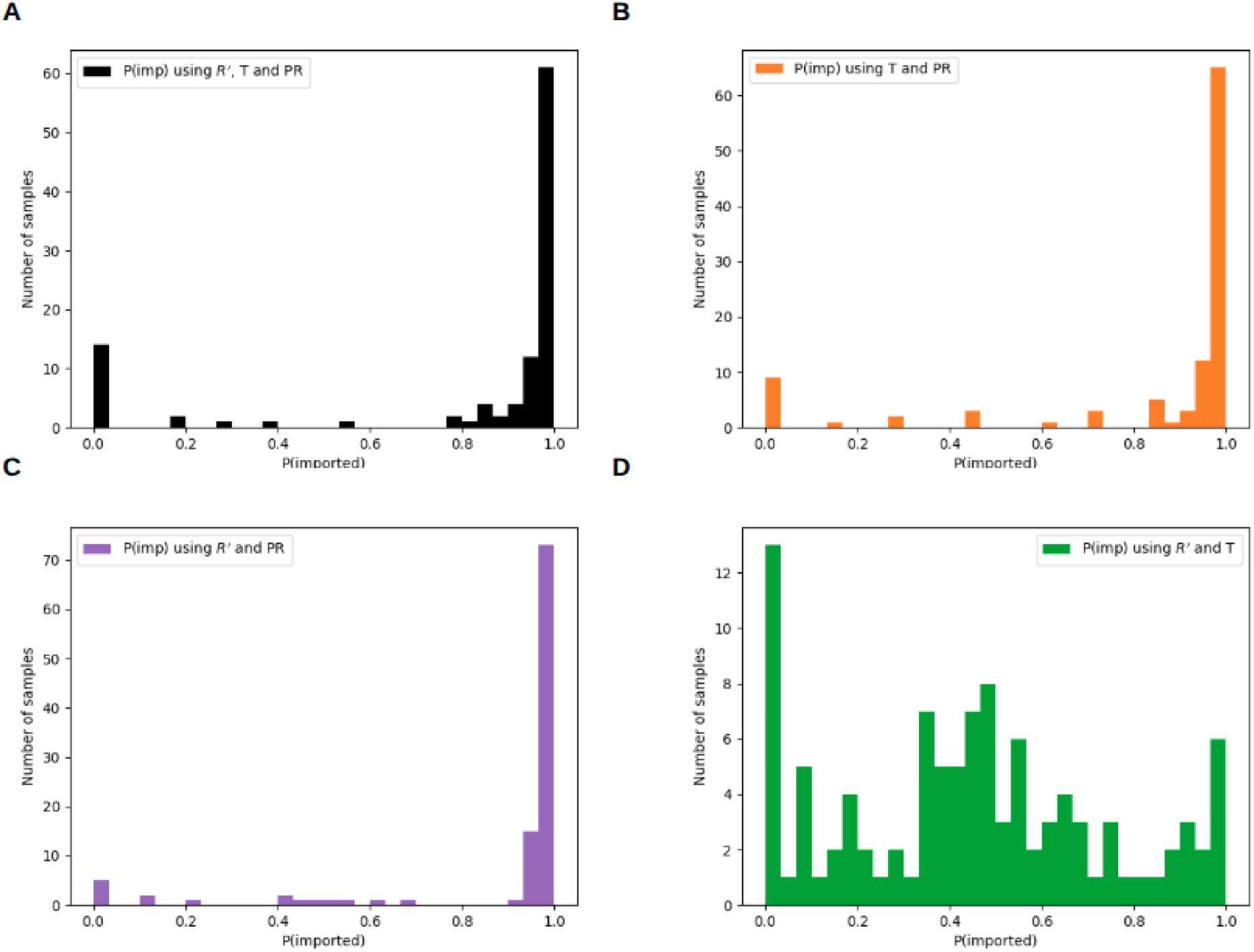
Distribution of importation probabilities using different combinations of factors. Probability distributions of the clinical cases with travel reports of being imported, depending on the inclusion of factors included in the estimation. The black histogram (**A**) shows the estimations from the formula described in Methods sections, while the orange (**B**), purple (**C**) and green (**D**) distribution show the estimations when removing the genetic relatedness (*R’*), the travel duration (T) or the transmission intensity (PR) metrics respectively.

**Appendix 1-Table 1:**
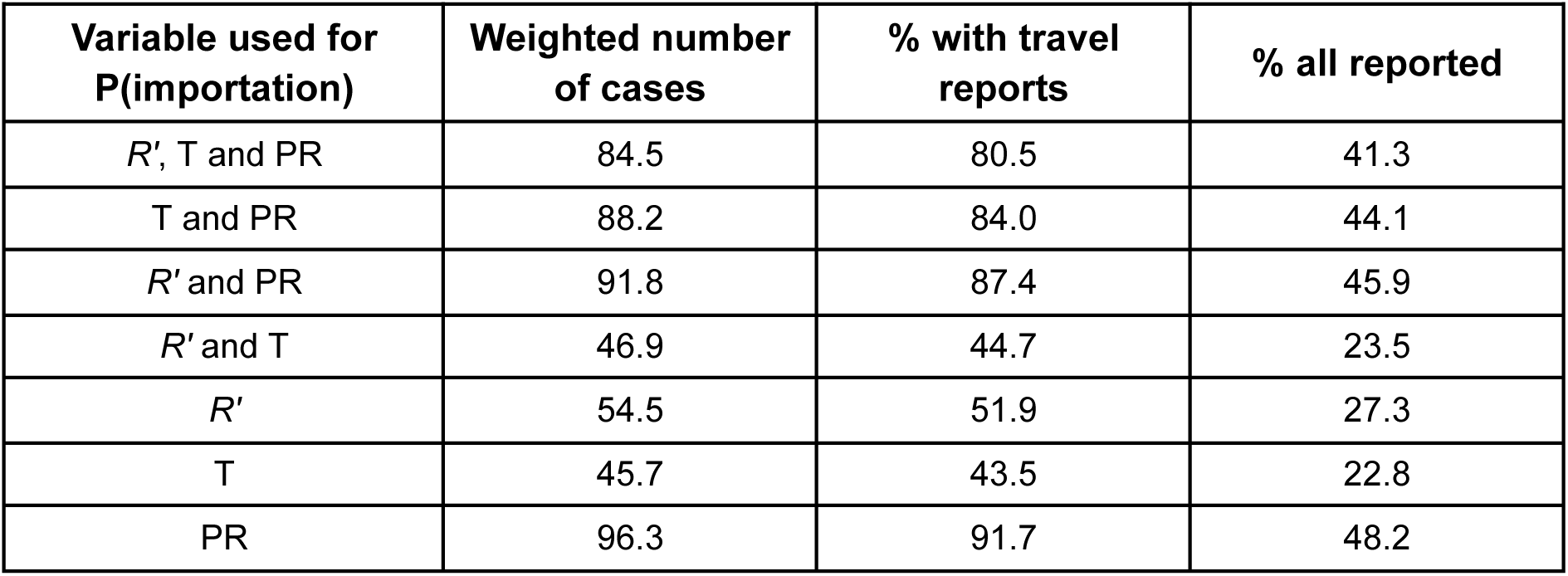
Importation probability statistics using different data sources in the estimations. The weighted number of cases (second column, n=2007) and their percentages with respect to all mRDT+ cases with travel reports (third column, n=1057) and with respect to all studied cases (last column, n=2007)) obtained for different combinations of data sources (specified in the first column). *R’*: the estimation of probability of importation (see Methods) includes the *R’* in the travel destination (fraction of samples from the travel destination province genetically related to the case) and origin (fraction of samples from the local province –Maputo-genetically related to the case). Otherwise, equal *R’* is assumed between origin and travel destination. T: the estimation of probability of importation includes the metrics of the travel duration and time spent at home. Otherwise, equal time is assumed. PR: the mRDT positivity rates (PR) per province in children reported in the last Health Demographic Survey 2022-2023 is used as a proxy of transmission intensity for each province. Otherwise, equal PR is assumed.

## Appendix 2 Supplementary tables

**Table 1:** Comparison of travel reports and genetic relatedness with Maputo province (local) and travel destination provinces. The table shows the number of cases that reported (Yes) or not (No) a travel to each province (columns “Destination province”) and how many of these cases show a fraction of genetically related pairs (IBD >0.1 with p<0.05) higher when compared with the samples collected in Maputo province (third column) or when compared with the travel destination province (fourth column) (n=200). The p-value shows the result of a Chi-square consistency test under the hypothesis that the genetic relatedness with Maputo or the other province does not depend on whether a travel has been reported or not.

| Destination province | Travel reported | More genetically related to Maputo | More genetically related to destination province | p-value |
| --- | --- | --- | --- | --- |
| Inhambane | No | 57 | 86 | 0.018 |
|  | Yes | 12 | 45 |  |
| Sofala | No | 107 | 92 | 1.000 |
|  | Yes | 1 | 0 |  |
| Manica | No | 119 | 80 | 0.846 |
|  | Yes | 0 | 1 |  |
| Zambézia | No | 114 | 71 | 0.171 |
|  | Yes | 6 | 9 |  |
| Nampula | No | 122 | 73 | 0.143 |
|  | Yes | 1 | 4 |  |
| Cabo Delgado | No | 103 | 96 | 0.976 |
|  | Yes | 0 | 1 |  |

**Table 2:** Factors associated with imported malaria among the 200 cases reported in Matutuine and Magude: distribution by malaria case classification (local vs imported) and odds ratios from univariate and multivariate logistic regression. SD=standard deviation; GM=geometric mean; CI=confidence interval.

| Category |  | Local | Imported | % imported | Univariate |  | Multivariate |  |
| --- | --- | --- | --- | --- | --- | --- | --- | --- |
|  |  |  |  |  | Odds ratio (95%CI) | p-value | Odds ratio (95%CI) | p-value |
| District of residence | Magude | 24 | 3 | 11.1% | 1 |  | 1 |  |
|  | Matutuine | 89 | 84 | 48.6% | 6.6 (2.3, 25.4) | <0.001 | 4.5 (1.5, 17.6) | 0.005 |
| Season | Dry | 29 | 35 | 54.7% | 1.9 (1.1, 3.5) | 0.029 | 1.6 (0.9, 3.0) | 0.147 |
|  | Rainy | 84 | 52 | 38.2% | 1 |  | 1 |  |
| Sex | Female | 51 | 50 | 49.5% | 1.6 (0.9, 2.9) | 0.085 | 1.5 (0.8, 2.9) | 0.194 |
|  | Male | 62 | 37 | 37.4% | 1 |  | 1 |  |
| Pregnant | Yes | 2 | 0 | 0.0% | 0.2 (0.0, 2.7) | 0.246 | NA |  |
|  | No | 35 | 34 | 49.3% | 1 |  | NA |  |
| Occupation | Domestic | 25 | 19 | 43.2% | 0.5 (0.2, 1.1) | 0.089 | 0.5 (0.2, 1.4) | 0.177 |
|  | Student | 17 | 8 | 32.0% | 0.3 (0.1, 0.8) | 0.019 | 0.4 (0.2, 1.2) | 0.094 |
|  | Farmer | 14 | 8 | 36.4% | 0.4 (0.1, 1.0) | 0.058 | 0.3 (0.1, 1.0) | 0.051 |
|  | Other | 37 | 21 | 36.2% | 0.4 (0.2, 0.8) | 0.011 | 0.4 (0.1, 1.4) | 0.164 |
|  | Unemployed | 20 | 31 | 60.8% | 1 |  | 1 |  |
| Age | Adult | 73 | 55 | 43.0% | 0.9 (0.5, 1.7) | 0.837 | 1.6 (0.7, 3.9) | 0.311 |
|  | Minor | 40 | 32 | 44.4% | 1 |  | 1 |  |
| Province travel destination | Gaza | 1 | 12 | 92.3% | 28.3 (3.9, 382.5) | <0.001 | NA |  |
|  | Inhambane | 3 | 54 | 94.7% | 52.9 (10.3, 390.8) | <0.001 | NA |  |
|  | Maputo | 8 | 2 | 20.0% | 1 |  | NA |  |
|  | Zambézia | 3 | 12 | 80.0% | 12.1 (2.2, 94.1) | 0.004 | NA |  |
|  | Manica | 0 | 1 | 100.0% | 10.2 (0.4, 1703.3) | 0.146 | NA |  |
|  | Maputo City | 2 | 0 | 0.0% | 0.7 (0.0, 13.2) | 0.668 | NA |  |
|  | Sofala | 0 | 1 | 100.0% | 10.2 (0.4, 1703.3) | 0.157 | NA |  |
|  | Nampula | 1 | 4 | 80.0% | 10.2 (1.2, 148.8) | 0.034 | NA |  |
|  | C. Delgado | 0 | 1 | 100.0% | 10.2 (0.4, 1703.3) | 0.150 | NA |  |
| Parasite | GM (SD) | 3.18 (1.1) | 3.08 (1.0) | NA | 0.9 (0.7, 1.2) | 0.529 | NA |  |
| **density** |  |  |  |  |  |  |  |  |
| **eCOI** | mean<br>(95%CI) | 1.53 (1.0,<br>3.4) | 1.80 (1.0,<br>4.2) | NA | 1.4 (1.0, 1.9) | 0.038 | NA |  |
| **Polyclon.** | Polyclonal | 54 | 50 | 48.1% | 1.5 (0.8, 2.6) | 0.176 | NA |  |
|  | Monoclonal | 59 | 37 | 38.5% | 1 |  | NA |  |
| **COI** | mean<br>(95%CI) | 2.21 (1.0,<br>5.2) | 2.80 (1.0,<br>7.8) | NA | 1.3 (1.1, 1.5) | 0.009 | NA |  |
| **prob.**<br>**polyclonal** | mean (SD) | 0.54 (0.5) | 0.65 (0.4) | NA | 1.8 (0.9, 3.3) | 0.075 | NA |  |
| **Naive**<br>**polyclon.** | Polyclonal | 67 | 62 | 48.1% | 1.7 (0.9, 3.1) | 0.081 | NA |  |
|  | Monoclonal | 46 | 25 | 35.2% | 1 |  | NA |  |

**Table 3:** Factor association to travel. Distribution of cases reporting and not reporting travel based on different factors and the statistical significance of the association of their odds ratio associated with travel. SD=standard deviation; GM=geometric mean; CI=confidence interval.

| Category |  | No travel | Travel | % travelled | Odds ratio (95%CI) | p-value | Multivariate |  |
| --- | --- | --- | --- | --- | --- | --- | --- | --- |
|  |  |  |  |  |  |  | Odds ratio (95%CI) | p-value |
| District of residence | Magude | 24 | 3 | 11.1% | 1 |  | 1 |  |
|  | Matutuine | 71 | 102 | 59.0% | 10.0 (3.5, 38.5) | <0.001 | 6.7 (2.3, 26.2) | 0.000 |
| Season | Dry | 23 | 41 | 64.1% | 2.0 (1.1, 3.7) | 0.025 | 1.6 (0.8, 3.0) | 0.178 |
|  | Rainy | 72 | 64 | 47.1% | 1 |  | 1 |  |
| Sex | Female | 43 | 58 | 57.4% | 1.5 (0.9, 2.6) | 0.161 | 1.3 (0.7, 2.4) | 0.487 |
|  | Male | 52 | 47 | 47.5% | 1 |  | 1 |  |
| Pregnant | Yes | 0 | 2 | 100.0% | 0.1 (0.0, 1.8) | 0.136 | NA |  |
|  | No | 41 | 28 | 40.6% | 1 |  | NA |  |
| Occupation | Domestic | 18 | 26 | 59.1% | 0.7 (0.3, 1.7) | 0.449 | 0.5 (0.2, 1.4) | 0.190 |
|  | Student | 16 | 9 | 36.0% | 0.3 (0.1, 0.8) | 0.012 | 0.4 (0.2, 1.2) | 0.099 |
|  | Farmer | 12 | 10 | 45.5% | 0.4 (0.2, 1.2) | 0.093 | 0.5 (0.2, 1.4) | 0.184 |
|  | Other | 32 | 26 | 44.8% | 0.4 (0.2, 0.9) | 0.023 | 0.4 (0.1, 1.4) | 0.151 |
|  | Unemployed | 17 | 34 | 66.7% | 1 |  | 1 |  |
| Age | Adult | 58 | 70 | 54.7% | 1.3 (0.7, 2.3) | 0.411 | 1.9 (0.8, 4.7) | 0.166 |
|  | Minor | 37 | 35 | 48.6% | 1 |  | 1 |  |
| Parasite density | GM (SD) | 3.18 (1.1) | 3.11 (1.0) | NA | 0.9 (0.7, 1.2) | 0.658 | NA |  |
| eCOI | mean (95%CI) | 1.48 (1.0, 3.6) | 1.80 (1.0, 4.1) | NA | 1.5 (1.1, 2.2) | 0.012 | NA |  |
| Polyclon. | Polyclonal | 63 | 41 | 39.4% | 2.0 (1.1, 3.5) | 0.018 | NA |  |
|  | Monoclonal | 42 | 54 | 56.3% | 1 |  | NA |  |
| COI | mean (95%CI) | 2.13 (1.0, 5.6) | 2.78 (1.0, 7.4) | NA | 1.3 (1.1, 1.6) | 0.003 | NA |  |
| prob. polyclonal | mean (SD) | 0.50 (0.5) | 0.67 (0.4) | NA | 2.4 (1.3, 4.4) | 0.007 | NA |  |
| Naive polyclon. | Polyclonal | 76 | 53 | 41.1% | 2.1 (1.2, 3.7) | 0.015 | NA |  |
|  | Monoclonal | 29 | 42 | 59.2% | 1 |  | NA |  |

**Table 4:**
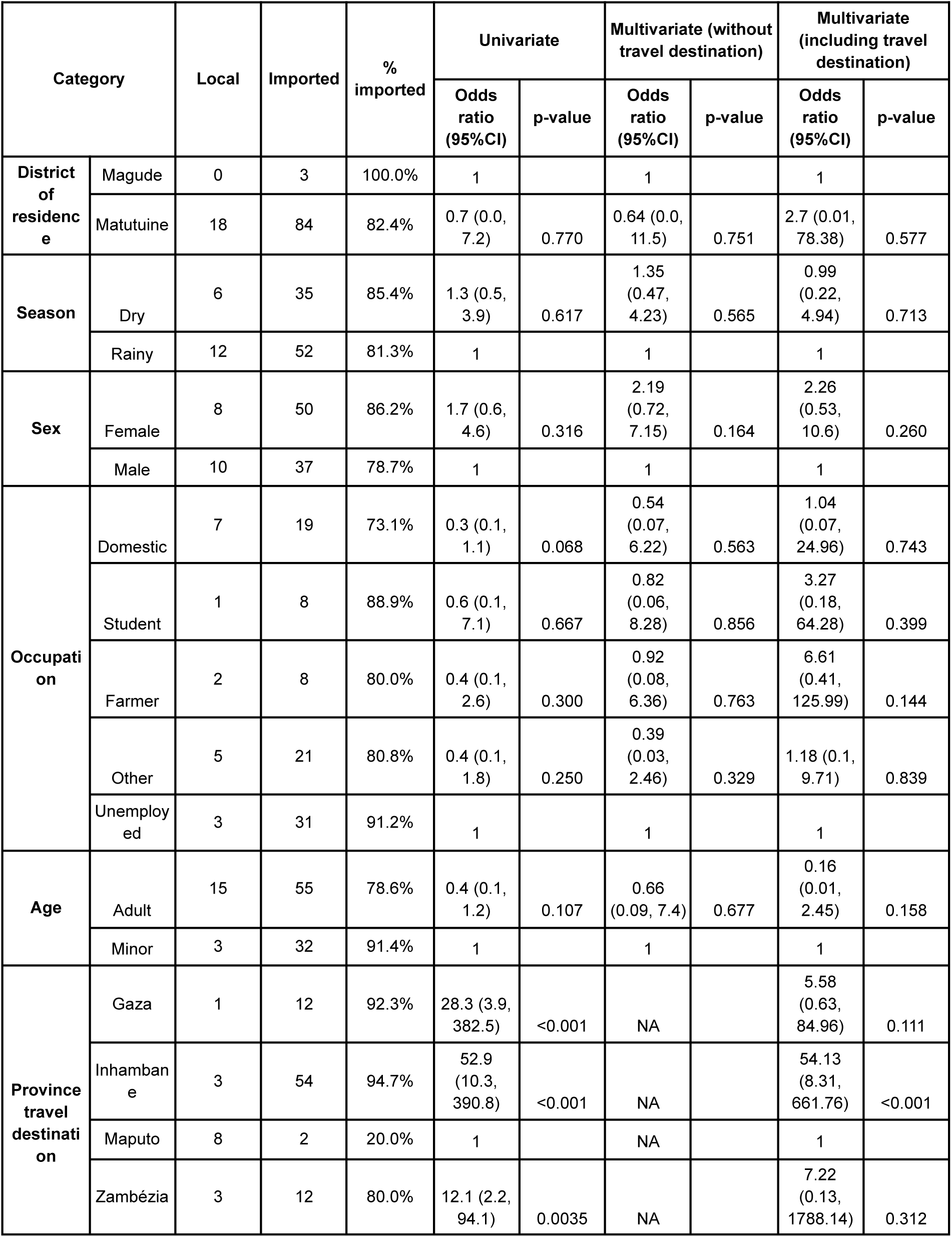
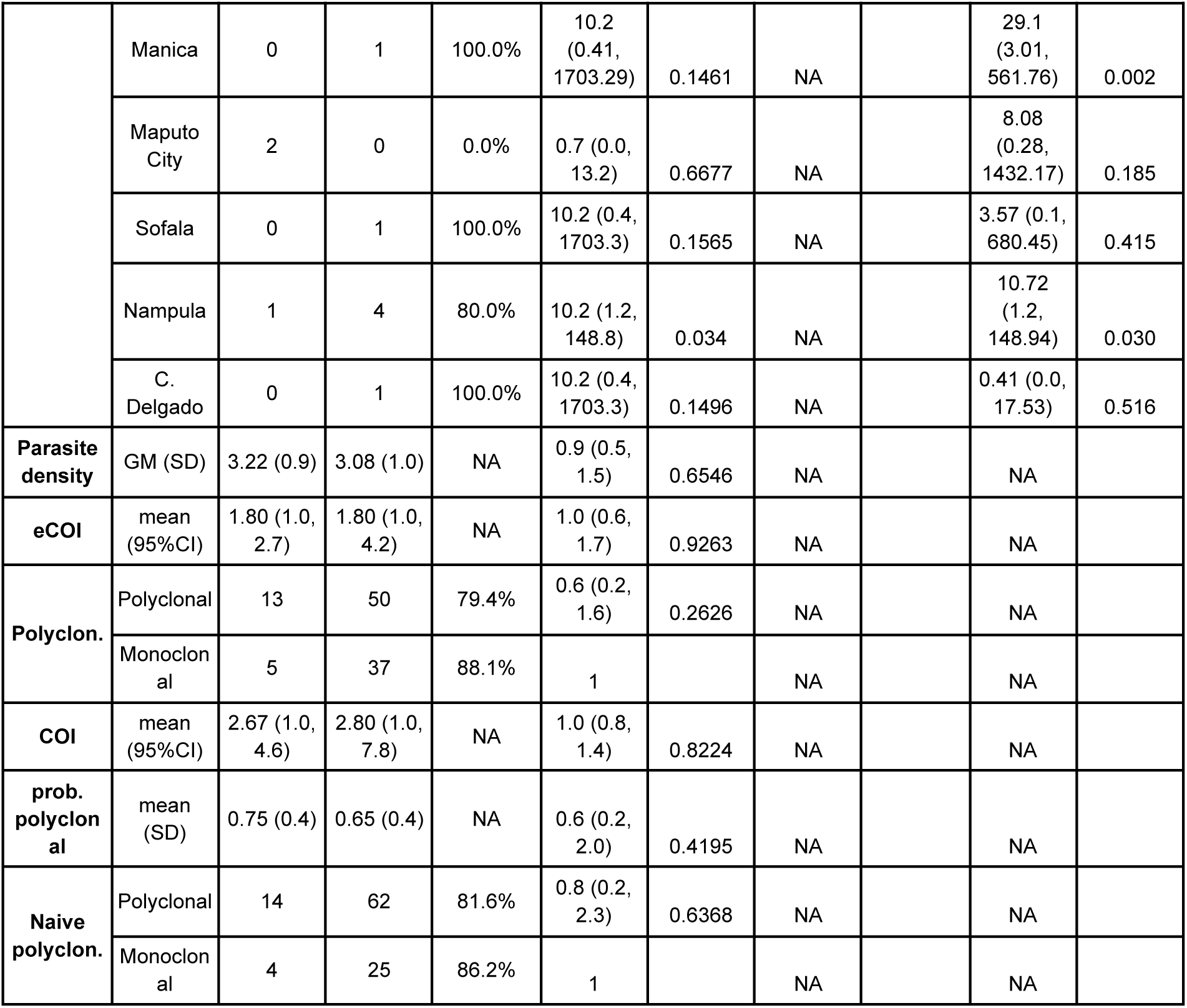
Factor association to importation conditioned to travel. Distribution of cases reporting travel based on different factors and the statistical significance of the association of their odds ratio associated with importation. SD=standard deviation; GM=geometric mean; CI=confidence interval.

## Notes

### Competing Interest Statement

The authors have declared no competing interest.

### Author Declarations

This study was conducted in accordance with the ethical principles outlined in the Declaration of Helsinki. Ethical approval was obtained from the Comite Nacional de Bioetica para Saude (CNBS) Mozambique, affiliated with the Ministerio de Saude (approval number 604/CNBS/21; 1st November 2023). The CNBS can be contacted at: Ministerio de Saude – 2 andar dto, Av. Eduardo Mondlane / Salvador Allende, PC 264, Maputo, Mozambique. I confirm that all necessary patient/participant consent has been obtained and the appropriate institutional forms have been archived, and that any patient/participant/sample identifiers included were not known to anyone (e.g., hospital staff, patients or participants themselves) outside the research group so cannot be used to identify individuals.

### Summary of Updates

Following the final round of revisions, we identified that a subset of samples included in the original analysis was affected by sampling errors. To identify potential issues of duplications in sample collection and library preparation, sample similarity was estimated using the root mean square error of within-sample allele frequency (WSAF) for highly diverse targets (pool D1.1), followed by clustering by Density-Based Spatial Clustering of Applications with Noise (DBSCAN) with ε=0.2. Clusters of samples (at least two samples) with clonality >2 and nearly identical WSAF across all loci, suggesting multiple DBSs were prepared using blood from a single patient, were excluded from the analysis. We therefore repeated the full analysis excluding these samples. The impact of this exclusion on the sample size per province was in general below 20%, with the exception of Nampula province, with a loss of approximately 50%. In addition, we applied more stringent quality-filtering criteria to ensure that the genetic analyses are not influenced by avoidable noise. These updates do not significantly change the results reported in the manuscript and do not alter the overall conclusions. As an example, the fraction of imported cases among the study samples, changed from 42.5% to 43.5%.

